# Changes in COVID-19-related outcomes and the impacts of the potential risk factors over time: a follow-up analysis

**DOI:** 10.1101/2021.01.02.21249140

**Authors:** Youfei Yu, Tian Gu, Thomas S. Valley, Lars G. Fritsche, Bhramar Mukherjee

**Author notes:** **Corresponding Author**: Bhramar Mukherjee, PhD, Department of Biostatistics, University of Michigan School of Public Health, 1415 Washington Heights, Ann Arbor, MI 48109.

## Abstract

**Importance:** Characteristics of COVID-19 patients changed over the course of the pandemic. Understanding how risk factors changed over time can enhance the coordination of healthcare resources and protect the vulnerable.

**Objective:** To investigate the overall trend of severe COVID-19-related outcomes over time since the start of the pandemic, and to evaluate whether the impacts of potential risk factors, such as race/ethnic groups, changed over time.

**Design:** This retrospective cohort study included patients tested or treated for COVID-19 at Michigan Medicine (MM) from March 10, 2020, to September 2, 2020. According to the quarter in which they first tested positive, the COVID-19-positive cohort were stratified into three groups: Q1, March 1, 2020 – March 31, 2020; Q2, April 1, 2020 – June 30, 2020; Q3, July 1, 2020 – September 2, 2020.

**Settings:** Large, academic medical center.

**Participants:** Individuals tested or treated for COVID-19.

**Exposure:** Examined potential risk factors included age, race/ethnicity, smoking status, alcohol consumption, comorbidities, body mass index (BMI), and residential-level socioeconomic characteristics.

**Main Outcomes and Measures:** The main outcomes included COVID-19-related hospitalization, intensive care unit (ICU) admission, and mortality, which were identified from the electronic health records from MM.

**Results:** The study cohort consisted of 53,853 patients tested or treated for COVID-19 at MM, with mean (SD) age of 44.8 (23.1), mean (SD) BMI of 29.1 (7.6), and 23,814 (44.2%) males. Among the 2,582 patients who tested positive, 719 (27.8%) were hospitalized, 377 (14.6%) were admitted to ICU, and 129 (5.0%) died. The overall COVID-positive hospitalization rate decreased from 41.5% in Q1 to 12.6% in Q3, and the overall ICU admission rate decreased from 24.5% to 5.3%. Black patients had significantly higher (unadjusted) overall hospitalization rate (265 [41.1%] vs 326 [23.2%]), ICU admission rate (139 [21.6%] vs 172 [12.2%]), and mortality rate (42 [6.5%] vs 56 [4.0%]) than White patients. Each quarter, the hospitalization rate remained higher for Black patients compared to White patients, but this difference was attenuated over time for the (unadjusted) odds ratios (Q1: OR=1.9, 95% CI [1.25, 2.90]; Q2: OR=1.42, 95% CI [1.02, 1.98]; Q3: OR=1.36, 95% CI [0.67, 2.65]). Similar decreasing patterns were observed for ICU admission and mortality. Adjusting for age, sex, socioeconomic status, and comorbidity score, the racial disparities in hospitalization between White and Black patients were not significant in each quarter of the year (Q1: OR=1.43, 95% CI [0.75, 2.71]; Q2: OR=1.25, 95% CI [0.79, 1.98]; Q3: OR=1.76 95% CI [0.81, 3.85]), in contrast to what was observed in the full cohort (OR=1.85, 95% CI [1.39, 2.47]). Additionally, significant association of hospitalization with living in densely populated area was identified in the first quarter (OR= 664, 95% CI [20.4, 21600]), but such association disappeared in the second and third quarters (Q2: OR= 1.72 95% CI [0.22, 13.5]; Q3: OR=3.69, 95% CI [0.103, 132]). Underlying liver diseases were positively associated with hospitalization in White patients (OR=1.60, 95% CI [1.01, 2.55], P=.046), but not in Black patients (OR=0.49, 95% CI [0.23, 1.06], P=.072, P_*int*_=.013). Similar results were obtained for the effect of liver diseases on ICU admission in White and Black patients (White: OR=1.75, 95% CI [1.01, 3.05], P=.047; Black: OR=0.46, 95% CI [0.17, 1.26], P=.130, P_*int*_=.030).

**Conclusions and Relevance:** These findings suggest that the COVID-19-related hospitalization, ICU admission, and mortality rates were decreasing over the course of the pandemic. Although racial disparities persisted, the magnitude of the differences in hospitalization and ICU admission rates diminished over time.

**Key Points:** *Questions:* How did the overall hospitalization and intensive care unit (ICU) admission rates change over the course of the pandemic and how did they vary by race?

*Findings:* In this cohort study of 2,582 patients testing positive for COVID-19, the unadjusted hospitalization rate decreased from 50.5% in Q1 (March 10, 2020, to March 31, 2020) to 17.9% in Q3 (July 1, 2020, to September 2, 2020) for Black patients, and from 23.2% in Q1 to 13.8% in Q3 for White patients. After adjusting for age, sex, sociodemographic factors, and comorbidity conditions, the odds ratios of hospitalization between White and Black patients were not significant in each quarter of the year 2020. No significant associations between ICU admission and race/ethnic groups were identified in each quarter or the entire three quarters.

*Meaning:* These findings suggests an appreciable decline in hospitalization and ICU admission rates among COVID-19 positive patients. The hospitalization and ICU admission rates across race/ethnic groups became closer over time.

## Introduction

Since the World Health Organization declared coronavirus disease 2019 (COVID-19) a pandemic on March 11, 2020,^1^ there have been over 77.1 million confirmed cases worldwide, including 1.70 million deaths in the following eight months.^2^ The United States has surpassed 10 million cases, with the state of Michigan topping 496,000 confirmed cases and over 12,000 deaths as of December 21, 2020.^3^ Studies find that racial and ethnic minority groups were disproportionately affected by COVID-19.^4–13^ For example, higher incidence of confirmed cases,^5,6,8,12^ increased risk of hospitalization^6,7,9,13^ and greater fatality burden^5,8,10,11^ were identified for Black individuals when compared to White individuals. Besides known racial health disparities that impact the severity of COVID-19 outcomes, our previous study found risk factors, such as cumulative comorbidity burden, to affect outcomes in White and Black patients differently.^9^

Emerging evidence shows an appreciable decline in mortality rate among patients with severe COVID-19 outcomes after the initial wave in March and April.^14–16^ A recent study conducted in a single health system in New York City found that the hospital mortality rate dropped from 25.6% at the start of the pandemic in March to 7.6% by mid-August 2020, adjusting for demographic and clinical factors.^15^ The same trend of improved survival over time was seen among COVID-19 patients requiring hospitalization^16^ or critical care management^14^ in England. Racial and ethnic disparities still remain, but recent data showed reduced differences in age-adjusted case fatality rate across ethnic groups.^17^ However, the dynamics of influencing risk factors on the change of COVID-19 outcomes over time remain widely unknown. Understanding how risk factors changed over the course of the pandemic can enhance the coordination of healthcare resources and protect the vulnerable.

As an follow-up of the work of Gu et al.,^9^ this study aims to examine the sociodemographic and clinical characteristics that are associated with various COVID-19 outcomes by race/ethnicity in an expanded COVID-19 cohort, as well as to characterize the risk factor trajectory over time, using electronic health records (EHRs) from Michigan Medicine (MM), a large academic health care system in the State of Michigan.

## Methods

### Study Cohort and COVID-19 Testing

The study sample consisted of 53,853 patients tested or treated for COVID-19 at MM, the University of Michigan Health System, from March 10, 2020, to September 2, 2020, with 2,582 testing positive. The sample was not randomly selected, as MM prioritized testing to those with COVID symptoms or at greatest risk of exposure, particularly during the early stages of the pandemic. Patients in the tested cohort received one of the four types of diagnostic tests: an in-house polymerase chain reaction (PCR) test (51,781 patients [96.2%]), a commercial antibody test (Viracor; 446 patients [.83%]), COVID-19 nasopharynx or oropharynx PCR tests deployed by the Michigan Department of Health and Human Services (54 patients [.10%]), reverse transcription–PCR tests performed in external labs (975 patients [1.8%]), and a small fraction of RNA test (4 patients [.007%]); 727 tested patients (1.3%) were transferred, tested elsewhere, or had no information on the type of testing they received.

### COVID-19 Outcomes

In this study, we focused on three different COVID-19-related outcomes among individuals who have tested positive: hospitalization, intensive unit care (ICU) admission, and mortality, which roughly represent various stages of disease progression. We also investigated the factors associated with being tested and testing positive for COVID-19 using a randomly selected untested comparison group (**eMethod** in the Supplement). The corresponding results are reported in the Supplement. The definitions of these COVID-19 outcomes are listed in **eTable 1**, and the sample sizes for each outcome are presented in **eFigure 1** in the Supplement.

### Definition of Demographics, Socioeconomic Status, Comorbidities, and Other Adjusted Covariates

We followed the same process described in Gu et al.^9^ to extract the relevant variables from the EHRs. The socio-demographic variables considered included age, self-reported sex, race/ethnicity, smoking status, alcohol consumption, body mass index (BMI), neighborhood socioeconomic disadvantage index (NDI),^18^ and population density (in persons per square mile).^18^ To characterize the underlying medical conditions of the patients, we considered seven comorbid conditions: respiratory conditions, circulatory conditions, any cancer, type 2 diabetes, kidney diseases, and autoimmune diseases, all of which were coded binary. The general health status of patients was represented by a comorbidity score ranging from 0 to 7 that sums up the aforementioned seven conditions. In addition, those who had at least one encounter in any of the primary care locations in MM since January 1, 2018, were regarded as seeking primary care.

### Statistical Analysis

For each COVID-related outcome *Y*_*COVID*_, we fit a Firth’s bias-corrected logistic regression to avoid the potential separation issues in the traditional maximum likelihood estimates. The analysis model is given by

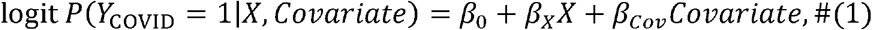

where *X* and *Covariate* denote the risk factor of interest and the vector of covariates, respectively. We explored four nested sets of variables for *Covariate*, respectively labeled adjustment 0-3 (defined in **eTable 1** in the Supplement), to check the robustness of inference to the choice of potential confounders (**eTable 2** in the Supplement). Our final model (adjustment 3) adjusted for age, sex, race/ethnicity, population density, NDI, and comorbidity score. We excluded the accumulative comorbidity score when the factor of interest was one of the individual comorbid conditions. The models for being tested and testing positive further adjusted for population density.

We further conducted a quarter-stratified analysis to assess the changes in racial disparities and the effects of other risk factors for severe COVID-19-related outcomes over time since the start of the pandemic. The Rac variable was self-reported and defined as four categories: White, Black, other known race/ethnicity, and unknown race/ethnicity. According to the quarter in which they first tested positive, the COVID-19-positive cohort were stratified into three groups: Q1, March 1, 2020 – March 31, 2020; Q2, April 1, 2020 – June 30, 2020; Q3, July 1, 2020 – September 2, 2020. Similarly, the COVID-19-negative cohort were stratified by the quarter in which they first received the tests. The logistic regression model (1) was fitted using data for each quarter.

To evaluate the effect of race/ethnicity on the potential risk factors, we fit a set of models with interactions by race:

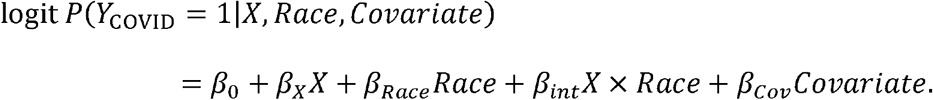

For each model, we report the Firth’s bias-corrected estimates of odds ratios along with their associated 95% Wald confidence intervals (CI) and P-values. For the interaction models, we also obtained the P-values of the differences in subgroup effects for White and Black patients by testing the null hypothesis *β*_*int*_ = 0. All analyses were performed using complete cases in R statistical software version 3.6.2 (R Project for Statistical Computing).

## Results

### Descriptive Statistics

**Table 1** summarizes the descriptive statistics for the tested cohort and each of the subgroups. The tested cohort included 53,853 patients, with mean (SD) age of 44.8 (23.1), mean (SD) BMI of 29.1 (7.6), and 23,814 (44.2%) males. During the study period, more White patients (n=38,977 [72.4%]) were tested than the other races/ethnicities, with 1,492 (66.1%), 17,213 (73.5%), and 19,911 (72.2%) White patients tested in Q1, Q2, and Q3, respectively. The proportions of patients of age:2:65 increased over time among those who have been tested (**eTables 3A-3C**). The untested comparison group consisted of 29,383 individuals, with mean (SD) age of 43.2 (24.4), mean (SD) BMI of 28.5 (7.3), and 13,581 (46.3%) males.

**Table 1.**
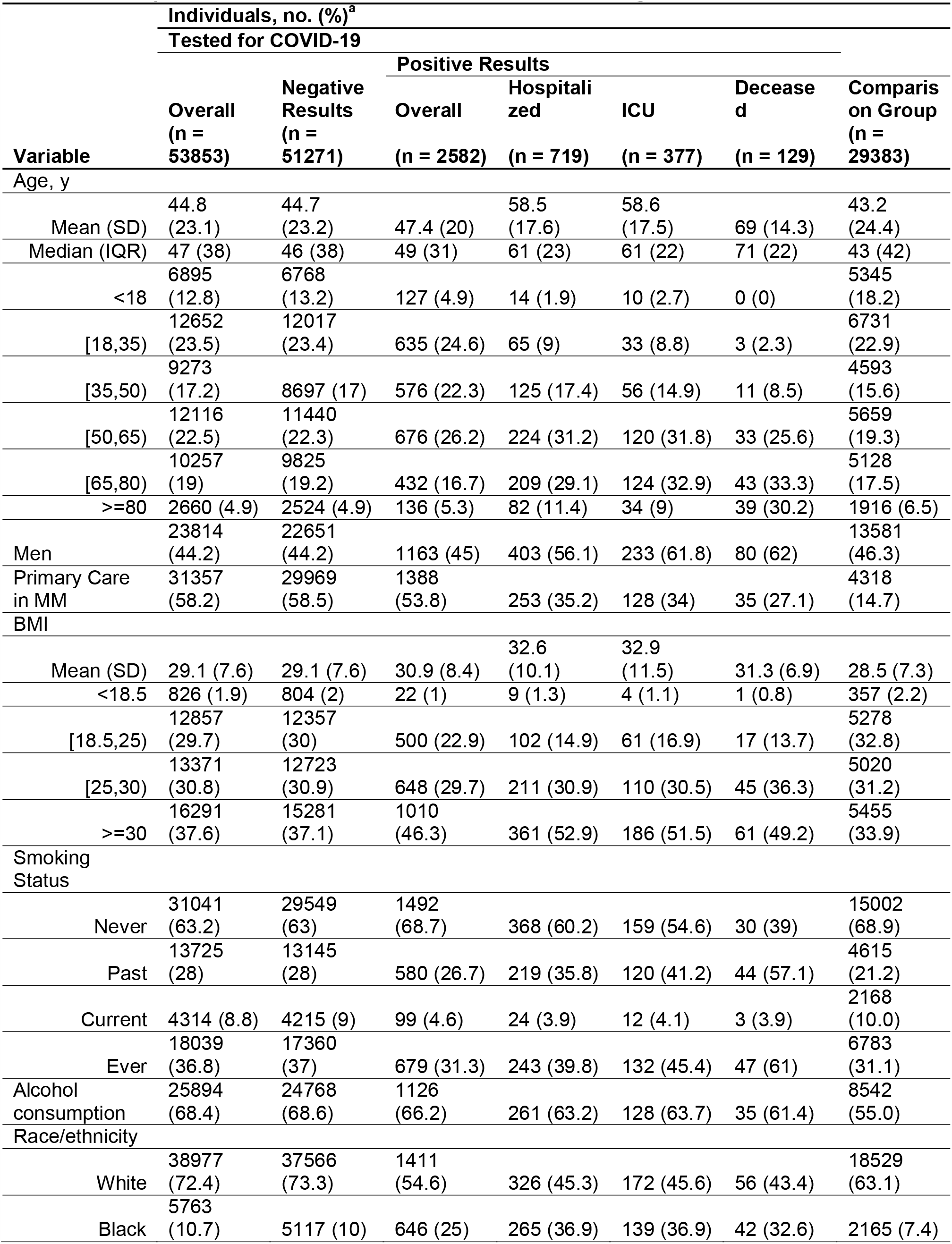
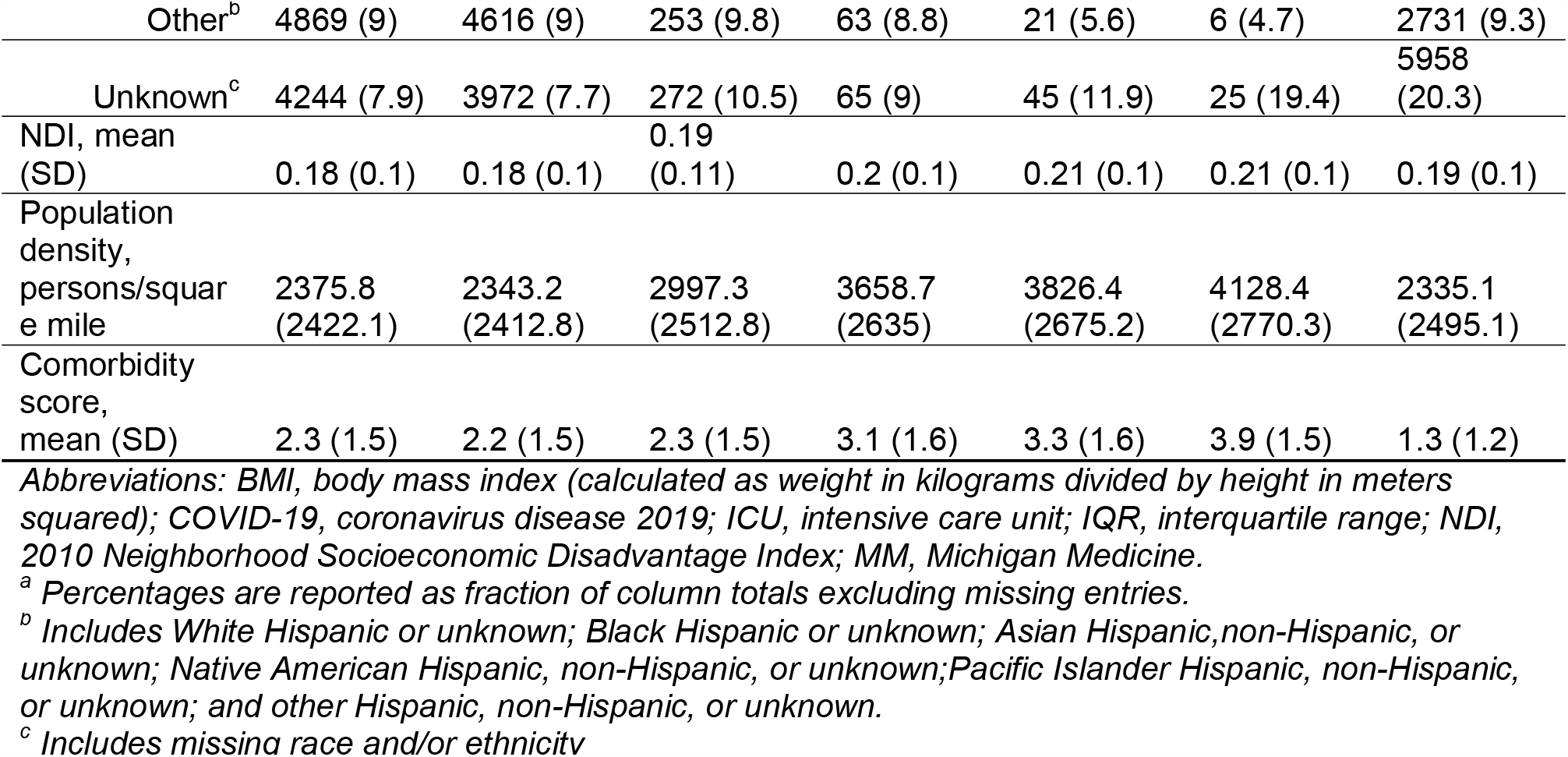
Descriptive Characteristics of the COVID-19 Tested or Diagnosed Cohort.

Among the 2,582 patients who have tested positive, 719 (27.8%) were hospitalized, 377 (14.6%) were admitted to ICU, and 129 (5.0%) died. Older patients, male sex, ever smokers, and increased comorbidity burden tended to be associated with worsened disease severity among COVID-19-positive patients. From Q1 to Q3, the mean and median age in general decreased over time for patients who were hospitalized or admitted to ICU. We observed a greater percentage of Black than White patients among those who were hospitalized or admitted to ICU patients in Q1. However, this relationship reversed in Q2 and Q3.

Stratifying the descriptive statistics by White and Black patients, we noted differences in covariate distributions between the two groups (**eTable 4**). For example, in the tested cohort, Black patients tended to be younger (mean [SD] age, 42.8 [21.7] vs 46.1 [23.3]), live in higher populated area (mean [SD] population density in persons per square mile, 3590 [2410] vs 2110 [2260]), have greater socioeconomic disadvantage (mean [SD] NDI, 0.18 [0.10] vs 0.09 [0.06]), and have higher burden of underlying medical conditions (mean [SD] comorbidity score, 2.53 [1.61] vs 2.29 [1.53]) than White patients. Similar pattern existed in the subgroups (e.g., hospitalized patients) of the tested cohort. The fractions of missingness for each characteristic are presented in **eTable 5**.

**Figure 1** indicated the changes in the overall and race-stratified COVID-19 outcomes over time. The overall hospitalization rate among patients testing positive decreased from 41.5% in Q1 to 12.6% in Q3, and the overall ICU admission rate decreased from 24.5% to 5.3%. In the full cohort, Black patients had significantly higher (unadjusted) hospitalization rate (265 [41.1%] vs 326 [23.2%]), ICU admission rate (139 [21.6%] vs 172 [12.2%]), and mortality rate (42 [6.5%] vs 56 [4.0%]) than White patients. When we examined each quarter, the hospitalization rate remained higher for Black patients compared to White patients, but this difference was attenuated over time for the (unadjusted) odds ratios (Q1: OR=1.9, 95% CI [1.25, 2.90]; Q2: OR=1.42, 95% CI [1.02, 1.98]; Q3: OR=1.36, 95% CI [0.67, 2.65]). Similar decreasing pattern was observed for ICU admission and mortality. In particular, Black patients had lower ICU admission rate in Q3 (4 [5.1%] vs 26 [5.8%]) and lower mortality rate in Q2 (15 [6.3%] vs 37 [9.2%]) and Q3 (0 [0%] vs 1 [0.2%]), though the differences were not statistically significant due to the small number of patients. At the end of the analysis, 22 patients were still hospitalized (n=5) or still in ICU (n=17).

**Figure 1.**
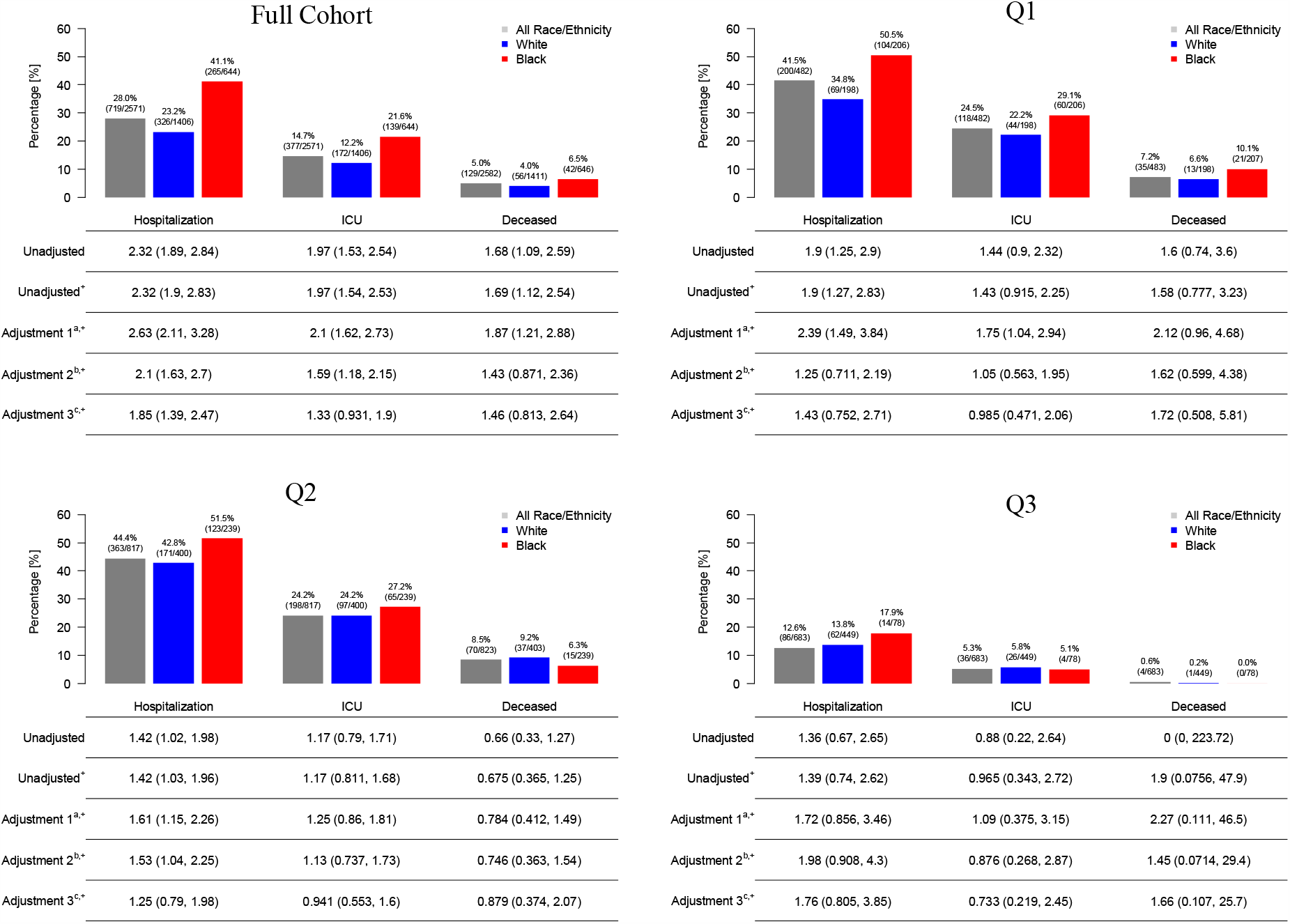
COVID-19 Outcomes Stratified by Race/Ethnicity in Each Quarter of Year 2020. Abbreviations: COVID-19, coronavirus disease 2019; ICU, intensive care unit; OR, odds ratio; Q1, March 10, 2020, to March 31, 2020; Q2, April 1, 2020, to June 30, 2020; Q3, July 1, 2020, to Sep 2, 2020. ^+^ Logistic regression with Firth’s correction. ^a^ Multivariable logistic regression with adjustment 1 (age + sex + race/ethnicity). Population density was also adjusted in the susceptibility (i.e. having positive test results) models. ^b^ Multivariable logistic regression with adjustment 2 (adjustment 1 + Neighborhood Socioeconomics Disadvantage Index). ^c^ Multivariable logistic regression with adjustment 3 (adjustment 2 + comorbidity score).

### Factors Associated with COVID-19 Testing

Overall, male sex, higher BMI, ever smoker, alcohol consumption, Black patients, lower NDI, areas with higher population densities, accumulation of comorbidities were associated with an increased chance of getting tested. These associations were fairly consistent over quarters (**eTable 6**).

### Factors Associated with COVID-19-Related Outcomes Among COVID-19-Positive Patients

Among the patients who tested positive for COVID-19 through September 2, 2020, Black patients had significantly higher (covariate adjusted) odds of being hospitalized compared to White patients (OR=1.85, 95% CI [1.39, 2.47]). The differences in ICU admission (OR=1.33, 95% CI [0.93, 1.90]) and mortality (OR=1.46, 95% CI [0.81, 2.64]) were not significant after covariate adjustment (**Figure 1**). Every 10-year increase in age, male sex, living in densely populated areas or disadvantaged neighborhoods, and heavier overall comorbidity burden were positively associated with hospitalization, ICU admission, and mortality (**Table 2**).

**Table 2.**
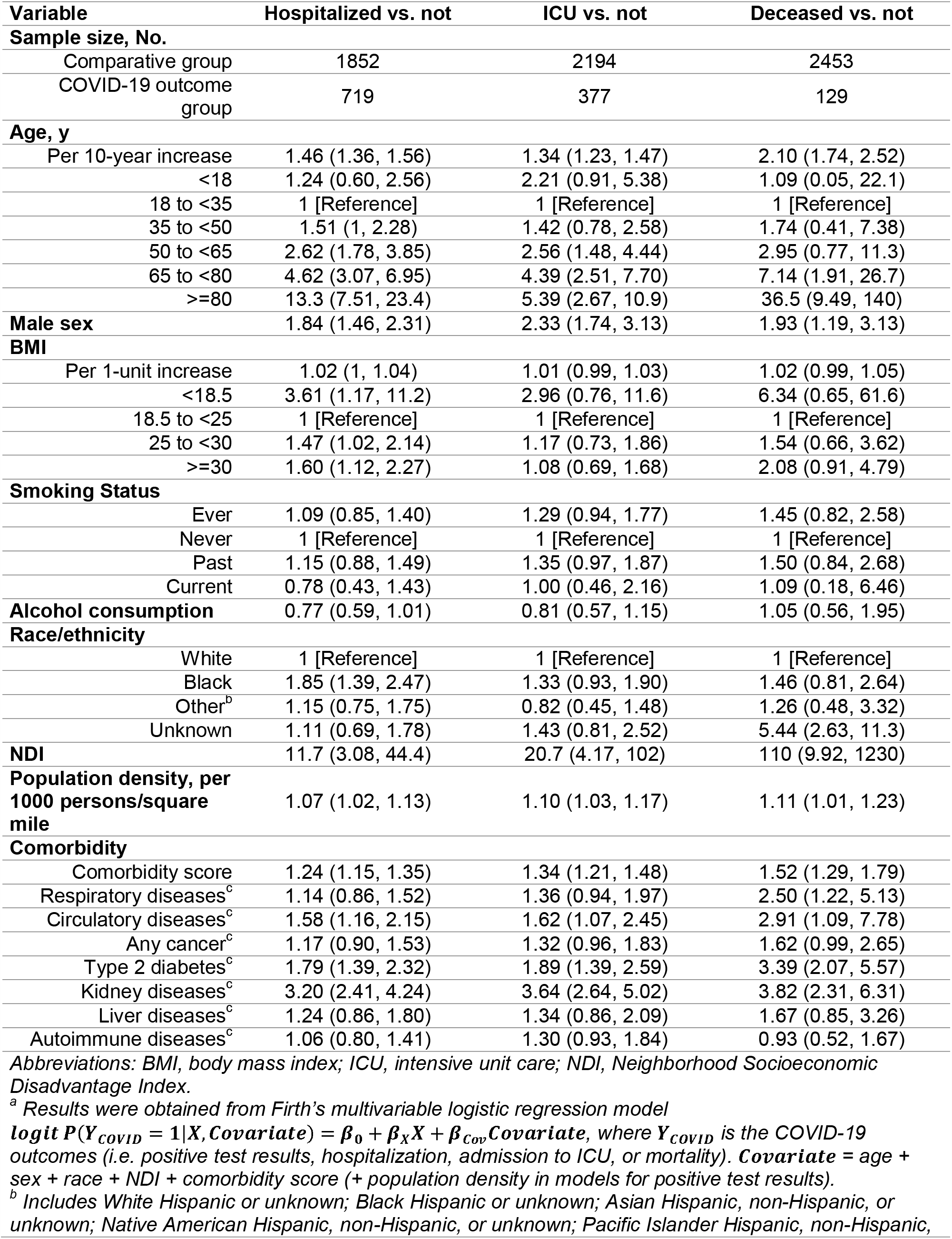
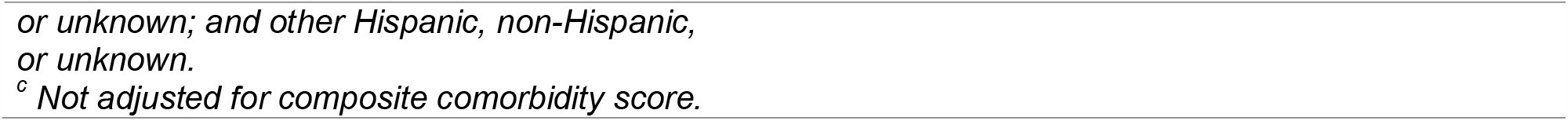
Odds Ratios (95% Confidence Intervals)^a^ for COVID-19-Related Outcomes in the Full Cohort.

| Variable | Hospitalized vs. not | ICU vs. not | Deceased vs. not |
| --- | --- | --- | --- |
| **Sample size, No.** |  |  |  |
| Comparative group | 1852 | 2194 | 2453 |
| COVID-19 outcome group | 719 | 377 | 129 |
| **Age, y** |  |  |  |
| Per 10-year increase | 1.46 (1.36, 1.56) | 1.34 (1.23, 1.47) | 2.10 (1.74, 2.52) |
| <18 | 1.24 (0.60, 2.56) | 2.21 (0.91, 5.38) | 1.09 (0.05, 22.1) |
| 18 to <35 | 1 [Reference] | 1 [Reference] | 1 [Reference] |
| 35 to <50 | 1.51 (1, 2.28) | 1.42 (0.78, 2.58) | 1.74 (0.41, 7.38) |
| 50 to <65 | 2.62 (1.78, 3.85) | 2.56 (1.48, 4.44) | 2.95 (0.77, 11.3) |
| 65 to <80 | 4.62 (3.07, 6.95) | 4.39 (2.51, 7.70) | 7.14 (1.91, 26.7) |
| >=80 | 13.3 (7.51, 23.4) | 5.39 (2.67, 10.9) | 36.5 (9.49, 140) |
| **Male sex** | 1.84 (1.46, 2.31) | 2.33 (1.74, 3.13) | 1.93 (1.19, 3.13) |
| **BMI** |  |  |  |
| Per 1-unit increase | 1.02 (1, 1.04) | 1.01 (0.99, 1.03) | 1.02 (0.99, 1.05) |
| <18.5 | 3.61 (1.17, 11.2) | 2.96 (0.76, 11.6) | 6.34 (0.65, 61.6) |
| 18.5 to <25 | 1 [Reference] | 1 [Reference] | 1 [Reference] |
| 25 to <30 | 1.47 (1.02, 2.14) | 1.17 (0.73, 1.86) | 1.54 (0.66, 3.62) |
| >=30 | 1.60 (1.12, 2.27) | 1.08 (0.69, 1.68) | 2.08 (0.91, 4.79) |
| **Smoking Status** |  |  |  |
| Ever | 1.09 (0.85, 1.40) | 1.29 (0.94, 1.77) | 1.45 (0.82, 2.58) |
| Never | 1 [Reference] | 1 [Reference] | 1 [Reference] |
| Past | 1.15 (0.88, 1.49) | 1.35 (0.97, 1.87) | 1.50 (0.84, 2.68) |
| Current | 0.78 (0.43, 1.43) | 1.00 (0.46, 2.16) | 1.09 (0.18, 6.46) |
| **Alcohol consumption** | 0.77 (0.59, 1.01) | 0.81 (0.57, 1.15) | 1.05 (0.56, 1.95) |
| **Race/ethnicity** |  |  |  |
| White | 1 [Reference] | 1 [Reference] | 1 [Reference] |
| Black | 1.85 (1.39, 2.47) | 1.33 (0.93, 1.90) | 1.46 (0.81, 2.64) |
| Other<sup>b</sup> | 1.15 (0.75, 1.75) | 0.82 (0.45, 1.48) | 1.26 (0.48, 3.32) |
| Unknown | 1.11 (0.69, 1.78) | 1.43 (0.81, 2.52) | 5.44 (2.63, 11.3) |
| **NDI** | 11.7 (3.08, 44.4) | 20.7 (4.17, 102) | 110 (9.92, 1230) |
| **Population density, per 1000 persons/square mile** | 1.07 (1.02, 1.13) | 1.10 (1.03, 1.17) | 1.11 (1.01, 1.23) |
| **Comorbidity** |  |  |  |
| Comorbidity score | 1.24 (1.15, 1.35) | 1.34 (1.21, 1.48) | 1.52 (1.29, 1.79) |
| Respiratory diseases<sup>c</sup> | 1.14 (0.86, 1.52) | 1.36 (0.94, 1.97) | 2.50 (1.22, 5.13) |
| Circulatory diseases<sup>c</sup> | 1.58 (1.16, 2.15) | 1.62 (1.07, 2.45) | 2.91 (1.09, 7.78) |
| Any cancer<sup>c</sup> | 1.17 (0.90, 1.53) | 1.32 (0.96, 1.83) | 1.62 (0.99, 2.65) |
| Type 2 diabetes<sup>c</sup> | 1.79 (1.39, 2.32) | 1.89 (1.39, 2.59) | 3.39 (2.07, 5.57) |
| Kidney diseases<sup>c</sup> | 3.20 (2.41, 4.24) | 3.64 (2.64, 5.02) | 3.82 (2.31, 6.31) |
| Liver diseases<sup>c</sup> | 1.24 (0.86, 1.80) | 1.34 (0.86, 2.09) | 1.67 (0.85, 3.26) |
| Autoimmune diseases<sup>c</sup> | 1.06 (0.80, 1.41) | 1.30 (0.93, 1.84) | 0.93 (0.52, 1.67) |

Abbreviations: BMI, body mass index; ICU, intensive unit care; NDI, Neighborhood Socioeconomic Disadvantage Index.
<sup>a</sup> Results were obtained from Firth's multivariable logistic regression model
$\text{logit } P(Y_{\text{COVID}} = 1 | X, \text{Covariate}) = \beta_0 + \beta_X X + \beta_{\text{Cov}} \text{Covariate}$ , where $Y_{\text{COVID}}$ is the COVID-19 outcomes (i.e. positive test results, hospitalization, admission to ICU, or mortality). $\text{Covariate}$ = age + sex + race + NDI + comorbidity score (+ population density in models for positive test results).
<sup>b</sup> Includes White Hispanic or unknown; Black Hispanic or unknown; Asian Hispanic, non-Hispanic, or unknown; Native American Hispanic, non-Hispanic, or unknown; Pacific Islander Hispanic, non-Hispanic,
*or unknown; and other Hispanic, non-Hispanic, or unknown.*
*<sup>c</sup> Not adjusted for composite comorbidity score.*

The racial disparities in hospitalization were not significant in each quarter of the year (Q1: OR=1.43, 95% CI [0.75, 2.71]; Q2: OR=1.25, 95% CI [0.79, 1.98]; Q3: OR=1.76 95% CI [0.81, 3.85]), in contrast to what was observed in the full cohort (**eTable 6**), possibly due to the reduced sample size after stratification. Additionally, significant association of hospitalization with living in densely populated areas was identified in the first quarter (OR= 664, 95% CI [20.4, 21600]), but such association disappeared in the second and third quarters (Q2: OR= 1.72 95% CI [0.22, 13.5]; Q3: OR=3.69, 95% CI [0.103, 132]). Similar pattern was observed for ICU admission (**eTable 6**).

Underlying liver diseases were positively associated with hospitalization in White patients (OR=1.60, 95% CI [1.01, 2.55], P=.046), but not in Black patients (OR=0.49, 95% CI [0.23, 1.06], P=.072, P_*int*_=.013) (**Figure 2A**). Similar results were obtained for the effect of liver diseases on ICU admission in White and Black patients (White: OR=1.75, 95% CI [1.01, 3.05], P=.047; Black: OR=0.46, 95% CI [0.17, 1.26], P=.130, P_*int*_=.030) (**Figure 2B**). Consistent with Gu et al.,^9^ the comorbidity score was a significant risk factor for hospitalization in White patients (OR=1.29, 95% CI [1.16, 1.44], P<0.001), but not in Black patients (OR=1.12, 95% CI [0.98, 1.29], P=0.098), though the interaction was not significant (P_*int*_=.15).

**Figure 2.**
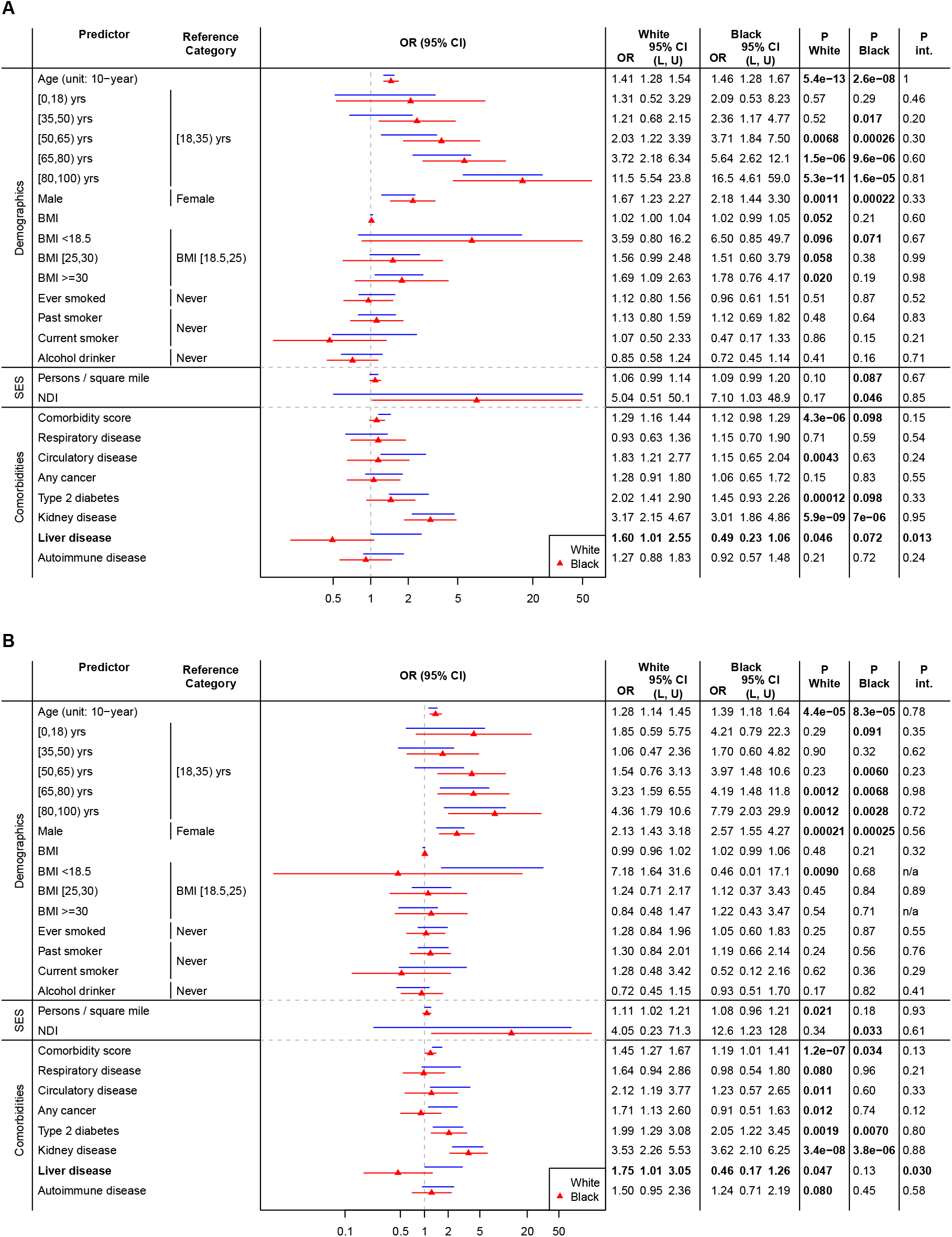
Hospitalization (A) and ICU Admission (B) for Black and White Patients in the Full Cohort. Abbreviations: BMI, body mass index; NDI, Neighborhood Socioeconomic Disadvantage Index. The results were from model *logit P(Y*_*COVID*_ = | *X, Covariate*) = *β*_0_ + *β*_*x*_ *X+ β*_*Race*_ *Race* + *β*_*int*_ *X × *Race + β*_COV_ Covariate*, where *Y*_*COVID*_ denotes hospitalization (A) or ICU admission (B), and *Covariate* =age + sex + NDI (+ comorbidity score in the demographic and socioeconomic status models). Results that are statistically significant at the level of 0.05 are bolded.

## Discussion

In this study, we evaluated the potential risk factors for COVID-19 prognosis using a larger cohort from the same healthcare system that covered a longer time frame than Gu et al.^9^ We observed that patients in racial minority group and with higher comorbidity burden were more susceptible to COVID-19 and more likely to be hospitalized, a result consistent with previous studies in the literature. On the other hand, the association between every 10-year increase in age and positive test results reversed compared to what was reported in Gu et al.,^9^ such that younger people, especially those aged 18-35 years, were more likely to be tested positive. The finding aligns with the recent data from Centers for Disease Control and Prevention, which shows an increase in the COVID-19 incidence in persons aged <30 years and a decrease in the median age for confirmed cases during May-August 2020.^19^

We observed a decline in the overall hospitalization and ICU admission rates over time, which may be partly explained by the rapid increase in testing from Q1 to Q3. The analysis stratified by quarters suggests less obvious contrast in COVID-19 outcomes between Black and White patients over time. Additionally, there was a reduction in socioeconomic disparity with regard to severe COVID-19 outcomes, as the socioeconomic variables NDI and population density were in general not significantly associated with hospitalization and ICU-admission in the second and the third quarter.

A large fraction (n=169 [23.5%]) of patients who were hospitalized in our sample was transferred from external hospitals, mainly from the Detroit Metro area. These transferred patients tended to have more severe symptoms and different distributions in sociodemographic characteristics from non-transferred patients. It is important to note the temporal changes in the context of care at MM (**eTable 7**). There was an enrichment in transferred patients in Q2 (n=93 [25.6%]) compared to Q1 (n=30 [15.0%]) and Q3 (n=11 [12.8%]) for those hospitalized. However, a sensitivity analysis that excluded patients who do not receive their primary care at Michigan Medicine was consistent with the overall analysis, namely improved outcomes and reduced health disparities over time (**eFigure 8**).

Our findings suggest that the COVID-19-related hospitalization, ICU admission, and mortality rates were decreasing over the course of the pandemic. Although racial disparities persisted, the magnitude of the differences in hospitalization and ICU admission rates diminished over time. This study also identifies a number of potential risk factors for COVID-19-related outcomes: older adults, male sex, racial minority groups, those with preexisting conditions, and those with poorer socioeconomic status. Our results can be useful for vaccine prioritization after the vaccines for COVID-19 becomes available.

## Data Availability

The data that support the findings of this study are available on request from the corresponding
author. The data are not publicly available due to privacy or ethical restrictions.

