## Supplement for "Changes in COVID-19-related outcomes and the impacts of the potential risk factors over time: a follow-up analysis"

**Supplemental Online Content**

**eMethods.** Analysis of the Susceptibility of COVID-19.

**eResults.** Factors associated with the Susceptibility of COVID-19.

**eFigure 1.** Flow Diagram of Patients Tested and Subsequent Patient Outcomes, Stratified by Race/Ethnicity.

**eFigure 2.** COVID-19 Testing and Susceptibility Stratified by Race/Ethnicity in Each Quarter of Year 2020.

**eFigure 3.** COVID-19 Susceptibility for White and Black Patients in the Full Cohort.

**eTable 1.** Sources of All Variables and Relevant Definitions

**eTable 2.** Odds Ratios of COVID-19 Outcomes From Logistic Regression for the Full Cohort.

**eTable 3.** Characteristics of the COVID-19 Tested or Diagnosed Cohort, Stratified by Quarters.

**eTable 4.** Descriptive Characteristics of the COVID-19 Tested or Diagnosed Cohort Stratified by White and Black Patients.

**eTable 5.** Missingness of the Variables in the Full Cohort, White, and Black Patients

**eTable 6.** Odds Ratios of COVID-19 Outcomes from Logistic Regression, Stratified by Quarters

**eTable 7.** Proportions of Transferred Patients by Outcome and Testing Quarter

**eTable 8.** Sensitivity Analysis Using Patients with Primary Care at Michigan Medicine, Stratified by Quarters.

**eMethods. Analysis of the Susceptibility of COVID-19**

**Selection of the Comparison Group**

In order to account for selection bias in our sample, we created a comparative group of untested individuals from the MM database. The comparative group included 29,383 randomly selected individuals who were alive on September 2, 2020 and have had at least one encounter with MM since April 23, 2020. The detailed inclusion and exclusion criteria are presented in eTable 1 in the Supplement.

**Statistical Methods**

We examined the characteristics associated with testing positive using the untested cohort described in the previous section for comparison. The analysis model is the same as Model (1) in the main text,$\begin{aligned} \mathrm{logit} P\left( \left. Y_{\mathrm{COVID}}=1 \right|X, Covariate \right)=\beta_{0}+\beta_{X}X+\beta_{Cov}Covariate, \end{aligned}$

where $X$ and $Covariate$ denote the risk factor of interest and the vector of covariates, respectively. Results are summarized in the Supplement (**eResults, eFigures 2 and 3, and eTable 6**).

**eResults. Factors associated with the Susceptibility of COVID-19.**

**eFigure 2** indicated the differences in COVID-19 susceptibility between White and Black patients. In the full cohort, the Black patients had significantly higher test positivity rate (646 [11.2%] vs 1411 [3.6%]) than White patients. When the cohort were stratified by quarter, the test positivity rate remained high for Black patients compared to White patients, but a decreasing trend over time was noted for the (unadjusted) odds ratios (Q1: OR=6.04, 95% CI [4.71, 7.75]; Q2: OR=3.86, 95% CI [3.26, 4.57]; Q3: OR=1.46, 95% CI [1.13, 1.87]).

In the full cohort, several factors were identified to be statistically different between the COVID-19-positive group and the untested controls. Specifically, Black patients had significant higher risk of being tested positive than White patients (OR=3.21, 95% CI [2.80, 3.67]) (**eTable 6**). Every 10-year increase in age was inversely associated with the odds of being tested positive (OR=0.97, 95% CI [0.94, 0.99]), as was male sex (OR=0.87, 95% CI [0.79, 0.96]) and ever smokers (OR=0.80, 95% CI [0.71, 0.90]). All of the comorbidity conditions considered were associated with an increased risk of having positive test results, with odds ratios ranging from 1.19 (95% CI [1.05, 1.34]) for any cancer to 3.74 (95% CI [3.34, 4.17]) for respiratory diseases.

The stratified analyses showed that most of the associations in each quarter stayed in the same direction as in the full cohort analysis (**eTable 6**). However, we observed a constant decrease over time in the odds of being tested positive for Black patients relative to White patients (Q1: OR=7.94, 95% CI [6.10, 10.3]; Q2: OR=3.50, 95% CI [2.79, 4.39]; Q3: OR=1.59, 95% CI [1.20, 2.12]).

Significant associations with positive test results were identified for overall comorbidity burden in both White (OR=1.53, 95% CI [1.47, 1.59], P<.001) and Black patients (OR=1.64, 95% CI [1.55, 1.75], P<.001), but it posed a higher risk on Black patients (P*_int_*=.001). Circulatory diseases, any cancer, type 2 diabetes, and kidney diseases showed similar directional results (**eFigure 2**).

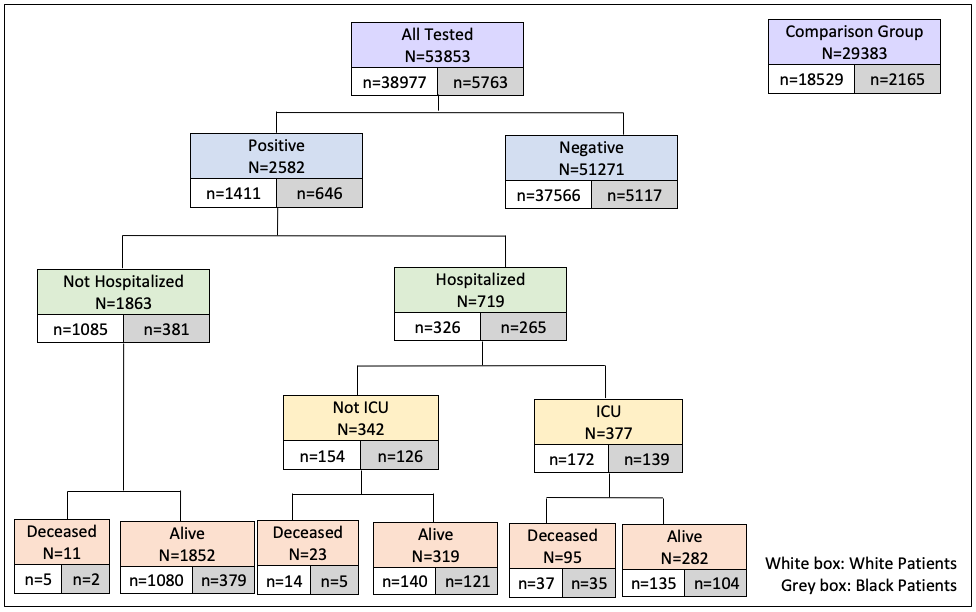

**eFigure 1. Flow Diagram of Patients Tested and Subsequent Patient Outcomes, Stratified by Race/Ethnicity**

For each COVID-19 outcome, we listed the total number of patients, as well as the total number of White patients (in the white box) and Black patients (in the grey box)

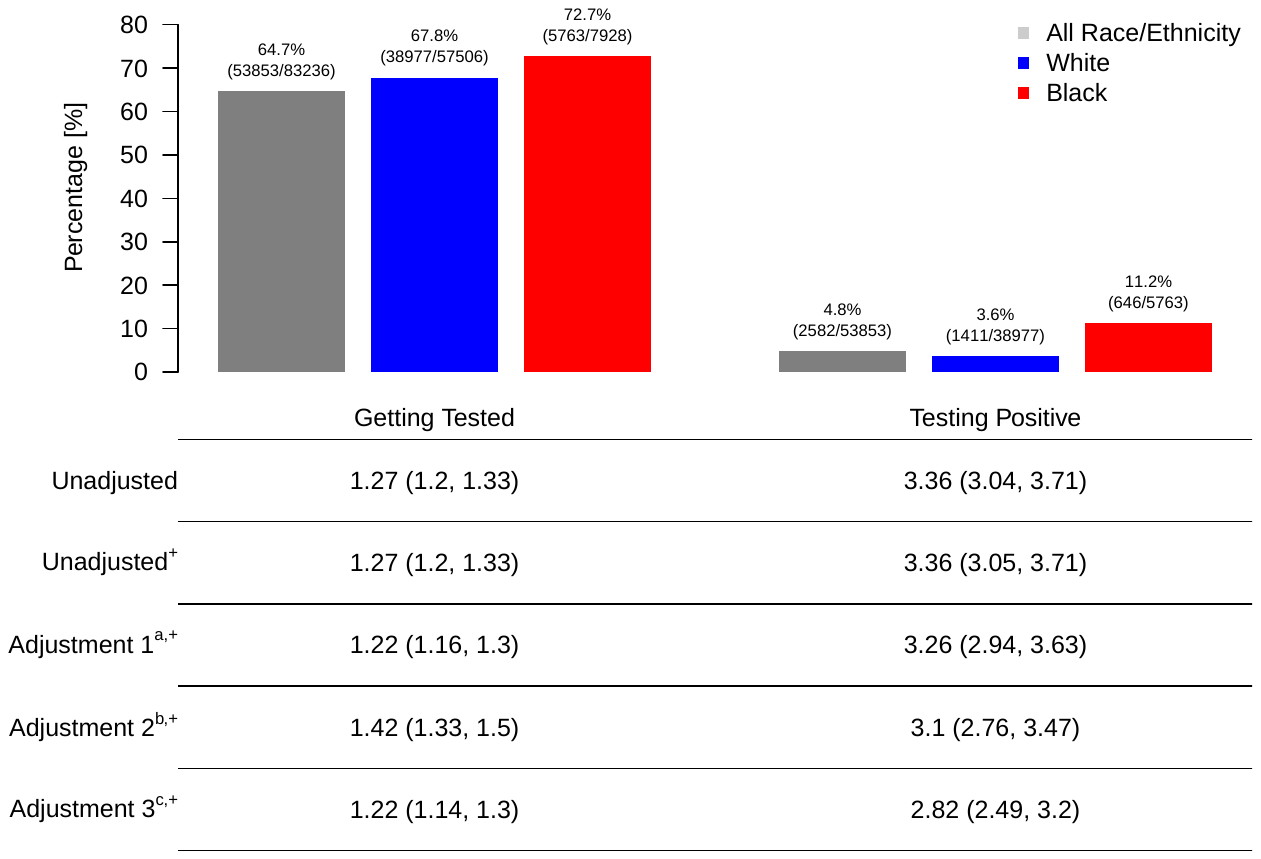

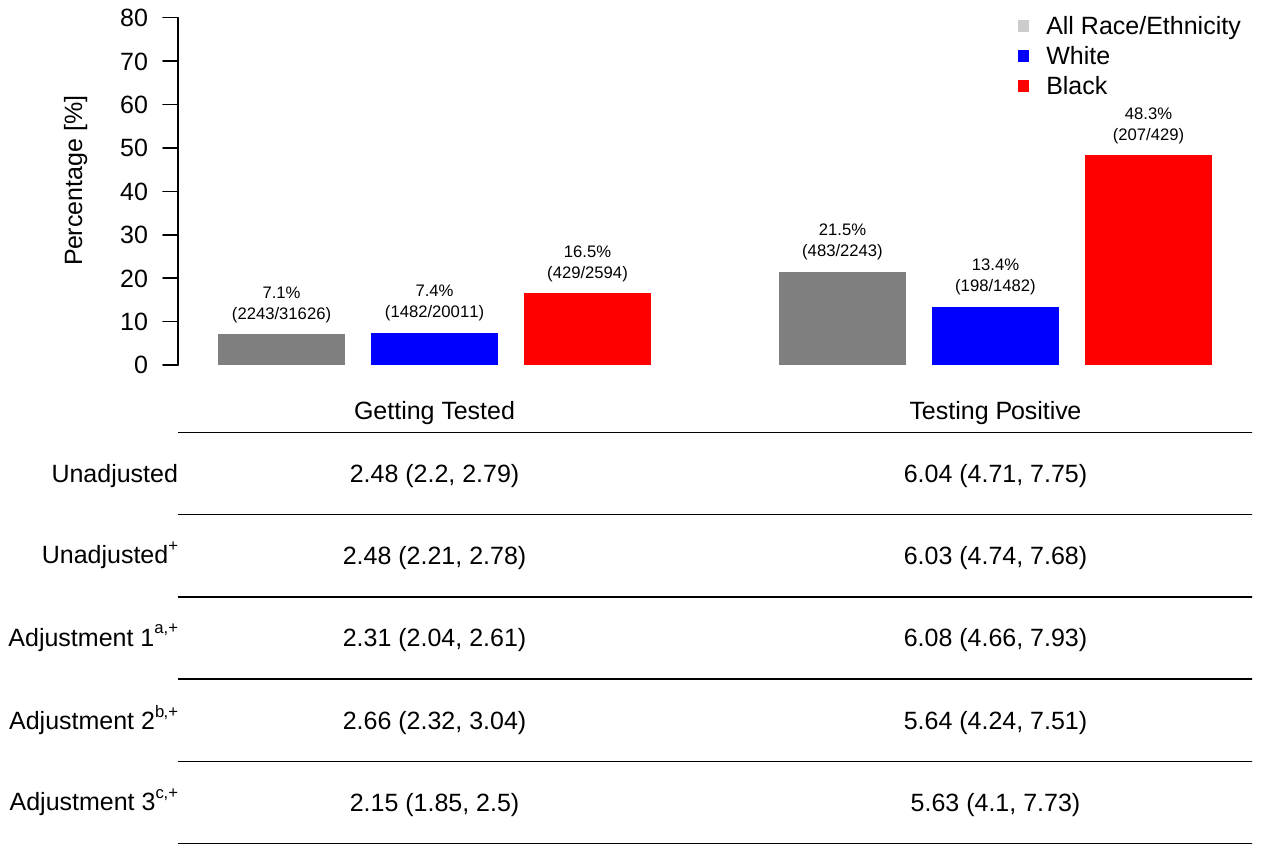

Q1

Full Cohort

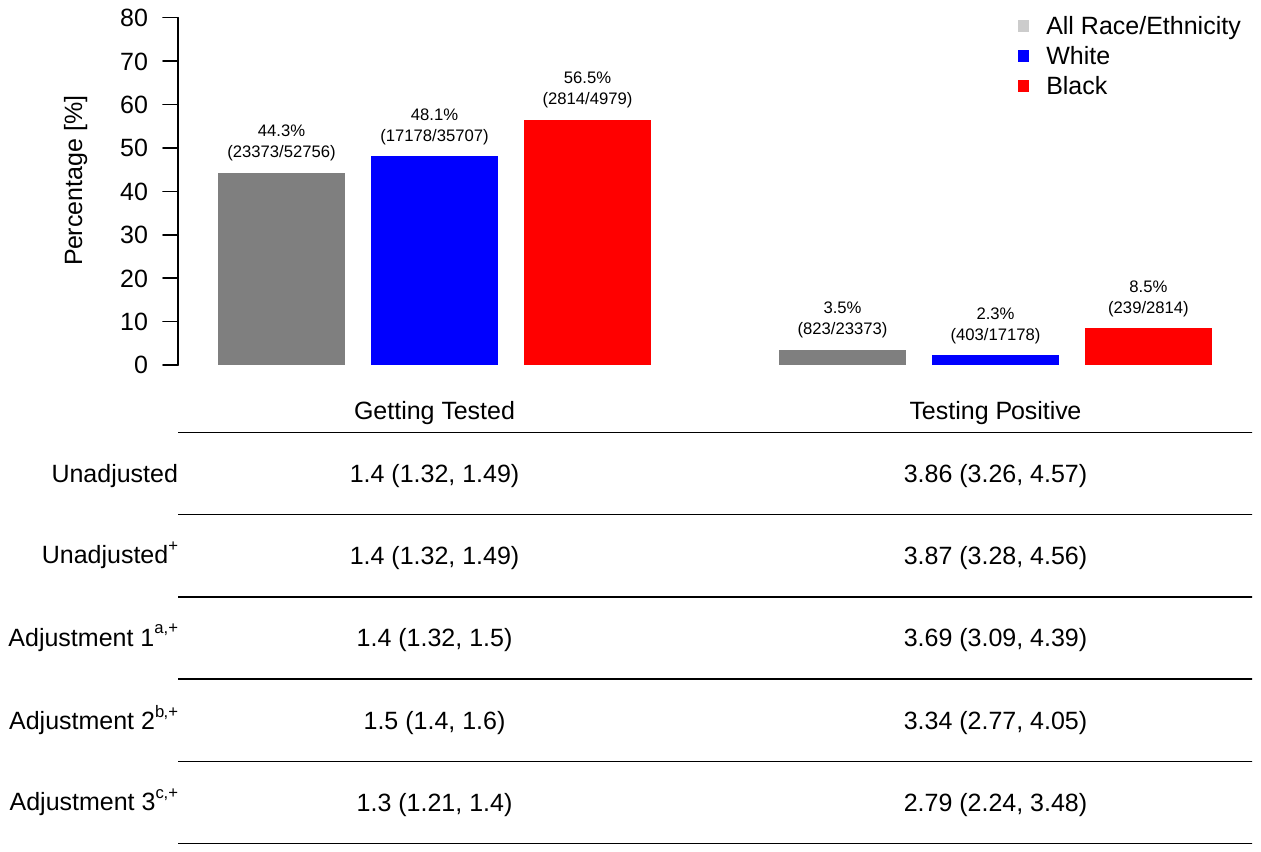

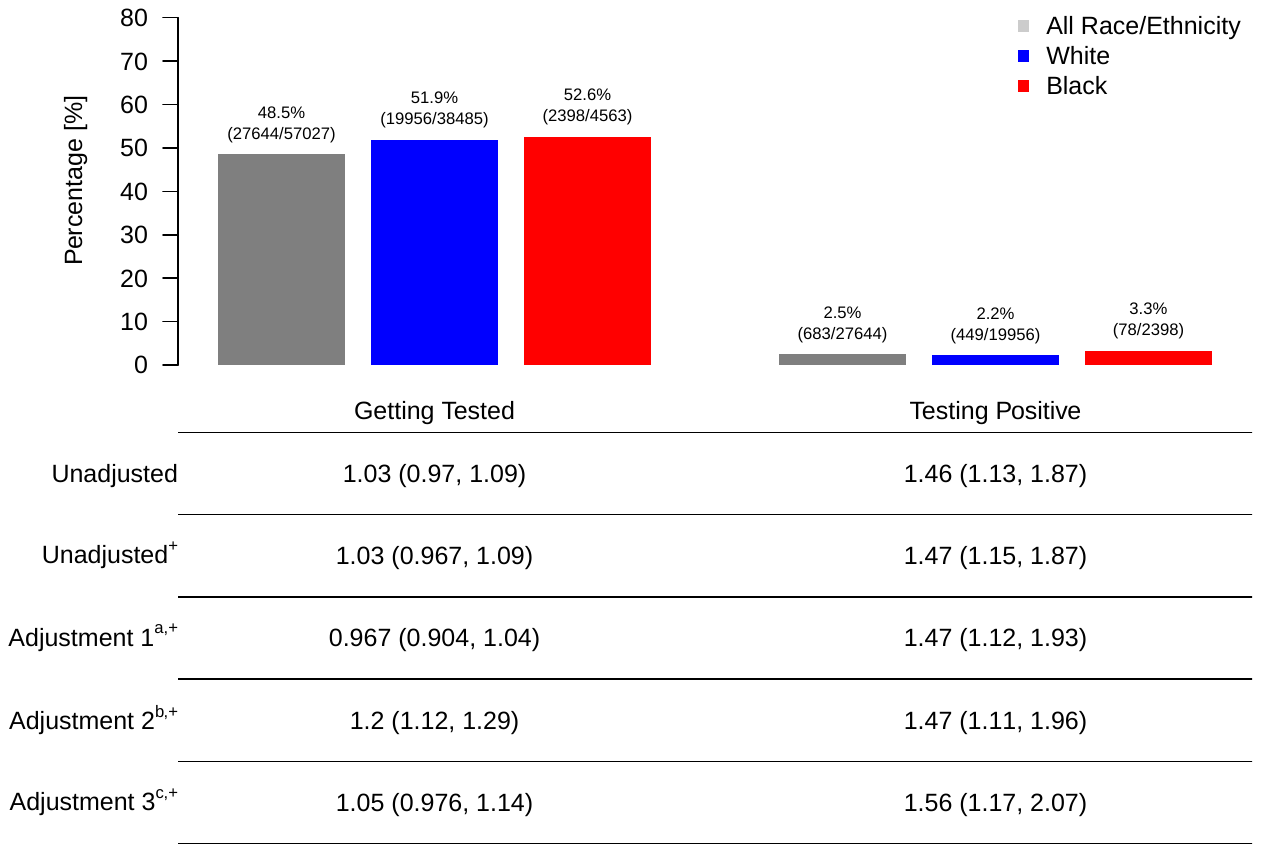

Q3

Q2

**eFigure 2. COVID-19 Testing and Susceptibility Stratified by Race/Ethnicity in Each Quarter of Year 2020.**

Abbreviations: COVID-19, coronavirus disease 2019; ICU, intensive care unit; OR, odds ratio; Q1, March 10, 2020, to March 31, 2020; Q2, April 1, 2020, to June 30, 2020; Q3, July 1, 2020, to Sep 2, 2020.

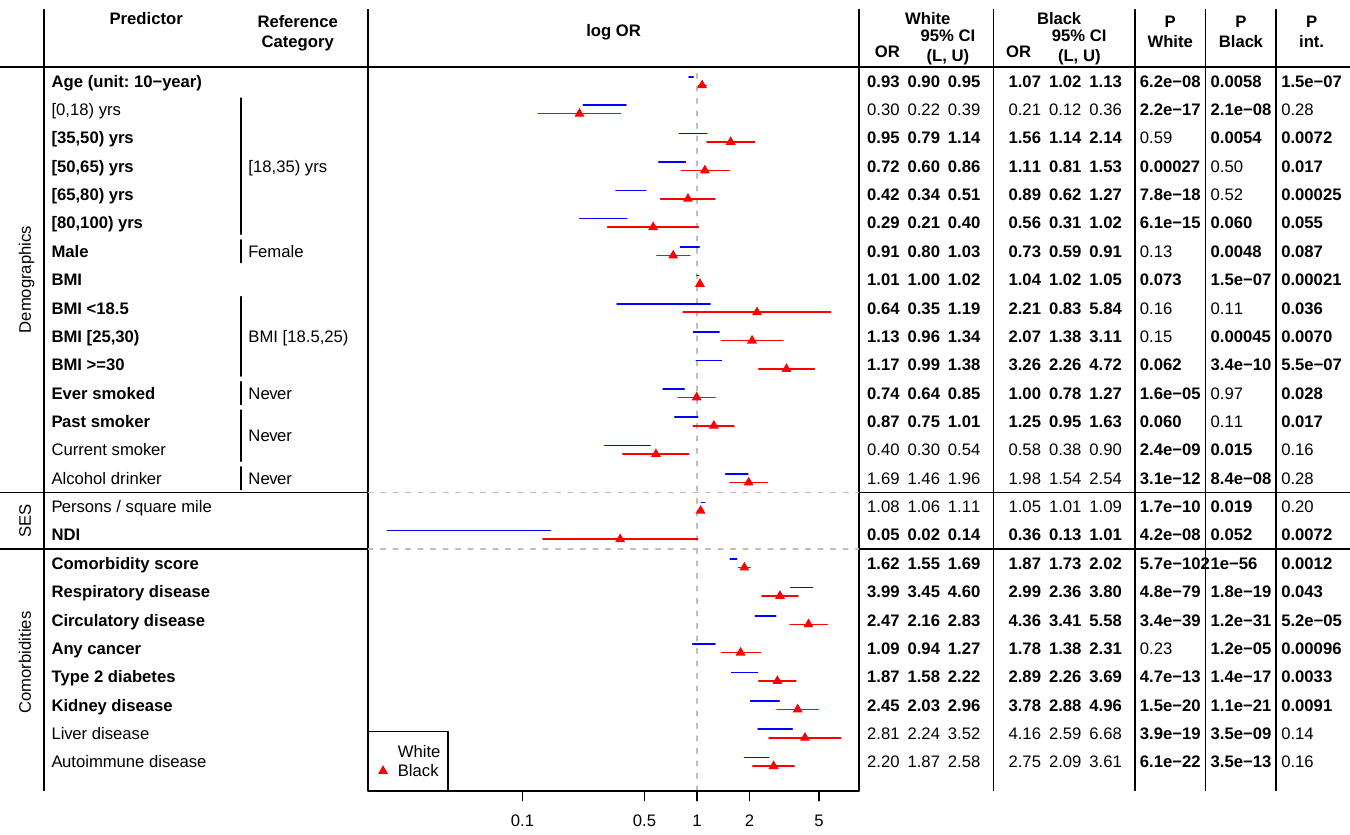

**eFigure 3. COVID-19 Susceptibility for White and Black Patients in the Full Cohort.**

Abbreviations: BMI, body mass index; NDI, Neighborhood Socioeconomic Disadvantage Index.

The results were from model $logit P\left( \left. Y_{COVID}=1 \right|X, Covariate \right)=\beta_{0}+\beta_{X}X+\beta_{Race}Race+\beta_{int}X\times Race+\beta_{Cov}Covariate$, where $Y_{COVID}$ denotes hospitalization (A) or ICU admission (B), and $Covariate$=age + sex + NDI + population density (+ comorbidity score in the demographic and socioeconomic status models). Results that are statistically significant at the level of 0.05 are bolded.

**eTable 1. Sources of All Variables and Relevant Definitions**

| **Variable** | **Definition** | **Sources** |
| --- | --- | --- |
| Age | Age of patient as of the data pull: 9/8/2020 | Electronic Health Record (EPIC) |
| Male | Gender of patient as reported. | Electronic Health Record (EPIC) |
| Primary Care in MM | If the patient has had an encounter in any of the primary care locations in MM since 01-01-2018, then 1; otherwise 0 | Derived from the Electronic Health Record |
| BMI | Excluded entries if (1) age at BMI measurement was missing or below 18 years, (2) height and/or weight were missing, (3) height measurements were below 69 cm or above 234 cm, (4) weight was above 400 kg, (5) BMI deviated more than one unit from BMI calculated from height and weight (BMI = weight [in kg] / height [in m]^2 ). Outliers for multiple values per person were defined as values that exceeded the median BMI +/- 3 x the median absolute deviation (MAD). Final BMI values was calculated as the median BMI of the remaining entries. | Derived from the Electronic Health Record |
| Ever-Smoker | If the last reported smoking status is "never", but reported smoking before 1; If the last reported smoking status is "never", and never reported smoking before, then 0 | Derived from the Electronic Health Record |
| **Smoking Status** |  | Self-Reported under Patient History in EHR |
| Never | If the patient never say that they are a 'former' smoker or 'current' smoker then 1, otherwise 0 |  |
| Past | If the last smoking status is "former", then 1 |  |
| Current | If the last smoking status is "current", then 1 |  |
| **Alcohol Consumption** | If the reported alcohol drinker status in the EHR was reported "yes" at least once; and never reported alcohol drinker before, then 0 | Self-Reported under Patient History in EHR |
| **Race/Ethnicity** |  | Patient Reported - Derived from the Electronic Health Record |
| White | If race was reported as "Caucasian" and ethnicity as "Hispanic or Latino" |  |
| Black | If race was reported as "African American" and ethnicity as "Hispanic or Latino" |  |
| Other / Known Ethnicity | If race was not reported as "African American" or "Caucasian"' and ethnicity was reported as "Non-Hispanic or Latino" or "Hispanic or Latino" |  |
| Other / Unknown Ethnicity | If race and/or ethnicity were missing |  |
| **SES** | Data defined by US census tract (based on residential address available in each patient’s EHR) for the year 2010 from the US Census and the American Community Survey (ACS). | The boundaries for the census tracts were normalized by 2010 tract boundaries using the Longitudinal Tract Data Base (Logan, Xu, and Stults, 2014). |
| NDI | 2010 Neighborhood Socioeconomic Disadvantage Index (without Proportion Black): mean of proportion of Population in Poverty; Unemployed; with Public Assistance Income; and Female-Headed Families with children. |  |
| Population density^a^ (1000-people/mi^2^) | Population density of the neighborhood that the patient lives in. |  |
| **Comorbidities** |  | Electronic Health Record (EPIC) |
| Respiratory Diseases | At least one of the following observed phecodes and their subcodes:  464, 465, 465.2, 465.4, 470, 471, 472, 473, 473.1, 473.3, 473.4, 474, 474.1, 474.2, 475, 475.9, 476, 477, 478, 479, 480, 480.1, 480.11, 480.12, 480.13, 480.2, 480.3, 480.5, 481, 483, 495, 495.1, 495.11, 495.2, 496, 496.1, 496.2, 496.21, 496.3, 497, 498, 499, 500, 500.1, 500.2, 501, 502, 503, 504, 504.1, 505, 506, 507, 508, 509, 509.1, 509.2, 509.3, 509.5, 509.8, 510, 510.2, 512, 512.1, 512.2, 512.3, 512.7, 512.8, 512.9, 513, 513.3, 513.31, 513.32, 513.4, 513.8, 514, 514.1, 514.2, 516, 516.1, 519, 519.1, 519.2, 519.8, 519.9 |  |
| Circulatory Diseases | At least one of the following observed phecodes and their subcodes:  394, 394.1, 394.2, 394.3, 394.4, 394.7, 395, 395.1, 395.2, 395.3, 395.4, 395.6, 396, 401, 401.1, 401.2, 401.21, 401.22, 401.3, 411, 411.1, 411.2, 411.3, 411.4, 411.41, 411.8, 411.9, 414, 414.2, 415, 415.1, 415.11, 415.2, 415.21, 416, 418, 418.1, 420, 420.1, 420.2, 420.21, 420.22, 420.3, 425, 425.1, 425.11, 425.12, 425.2, 425.8, 426, 426.2, 426.21, 426.22, 426.23, 426.24, 426.25, 426.3, 426.31, 426.32, 426.4, 426.7, 426.8, 426.9, 426.91, 426.92, 427, 427.1, 427.11, 427.12, 427.2, 427.21, 427.22, 427.3, 427.4, 427.41, 427.42, 427.5, 427.6, 427.61, 427.7, 427.8, 427.9, 428, 428.1, 428.2, 428.3, 428.4, 429, 429.1, 429.2, 429.3, 429.9, 430, 430.1, 430.2, 430.3, 433, 433.1, 433.11, 433.12, 433.2, 433.21, 433.3, 433.31, 433.32, 433.5, 433.6, 433.8, 440, 440.1, 440.2, 440.21, 440.22, 440.9, 441, 441.1, 441.2, 442, 442.1, 442.11, 442.2, 442.3, 442.4, 442.8, 443, 443.1, 443.7, 443.8, 443.9, 444, 444.1, 444.2, 444.5, 446, 446.1, 446.2, 446.3, 446.4, 446.5, 446.6, 446.7, 446.8, 446.9, 447, 447.1, 447.7, 448, 450, 451, 451.2, 452, 452.1, 452.2, 452.8, 453, 454, 454.1, 454.11, 455, 456, 457, 457.2, 457.3, 458, 458.1, 458.2, 458.9, 459, 459.1, 459.7, 459.9 |  |
| Any Cancer | At least one of the following observed phecodes and their subcodes:  145, 145.2, 145.3, 145.4, 149, 149.1, 149.2, 149.3, 149.4, 149.5, 149.9, 150, 151, 153, 153.2, 153.3, 155, 155.1, 157, 158, 159, 159.2, 159.3, 159.4, 164, 165, 165.1, 170, 170.1, 170.2, 172, 172.1, 172.11, 172.2, 172.21, 172.22, 172.3, 174, 174.1, 174.11, 175, 180, 180.1, 180.3, 182, 184, 184.1, 184.11, 184.2, 185, 187, 187.1, 187.2, 189, 189.1, 189.11, 189.12, 189.2, 189.21, 189.4, 190, 191, 191.1, 191.11, 193, 194, 195, 195.1, 195.3, 196, 197, 198, 198.1, 198.2, 198.3, 198.4, 198.5, 198.6, 198.7, 199.4, 200, 200.1, 201, 202, 202.2, 202.21, 202.22, 202.23, 202.24, 204, 204.1, 204.11, 204.12, 204.2, 204.21, 204.22, 204.3, 204.4, 209 |  |
| Type 2 Diabetes | At least one of the following observed phecode and their subcodes: 250.2 |  |
| Kidney Diseases | At least one of the following observed phecodes and their subcodes: 585 |  |
| Liver Diseases | At least one of the following observed phecodes and their subcodes: 571 |  |
| Autoimmune Diseases | At least one of the following observed phecodes and their subcodes: 242.1, 250.1, 335, 557.1, 694.1, 695.4, 696.4, 697, 704.1, 714.1, 717 |  |
| Comorbidity Score | The summation of 7 comorbidities values above, ranging from 0 to 7 |  |
| **COVID-19 Outcomes** |  | Derived from RDW's COVID Registry |
| COVID-19 Tested | Patients who were tested for COVID-19 at the time of data pull. |  |
| COVID-19 Positive | Patients who tested positive at least once for COVID-19 |  |
| COVID-19 Negative | Patients who always tested negative for COVID-19 |  |
| Non-Hospitalized | Patients in the positive COVID cohort who have no inpatient stays after 3/5/2020 |  |
| Hospitalized | Patients in the positive COVID cohort who checked in as an inpatient after 3/5/2020 at least once. |  |
| ICU | Patients in the positive COVID cohort who checked into the ICU during their inpatient stay after 3/5/2020. |  |
| Deceased | Patients in the cohort who have died based on their Electronic Health Record. | Electronic Health Record (EPIC) |
| **Comparison Group** | Randomly picked cohort of patients who are not part of the COVID-19 cohort (Tested, Positive, Negative), who are alive and who have had an encounter in MM (Inpatient, Outpatient or Emergency) since 2012-04-23. We created an untested comparison group (n=29,383) from the MM database, which is a similar-sized random sample of contemporaneous patients. Specifically, we initially extracted 30,000 individuals before limiting the group to patients who (1) were alive at the time of data pull, (2) have had encounters after 2012-04-22 and (3) who had inpatient, outpatient and/or emergency visits. At the time of the last update for COVID-19 outcomes (September 2, 2020), all patients in the comparison group were alive. | Derived from the Electronic Health Record |
| **Adjustments** |  |  |
| Adjustment 0 | Unadjusted |  |
| Adjustment 1 | age + sex + race/ethnicity (+ population density)* |  |
| Adjustment 2 | adjustment 1 + NDI |  |
| Adjustment 3 | adjustment 2 + comorbidity score |  |
| **Quarters** |  |  |
| Quarter 1 | March 1—March 31, 2020 |  |
| Quarter 2 | April 1 – June 30, 2020 |  |
| Quarter 3 | July 1—September 2, 2020 |  |

Abbreviations: MM, Michigan Medicine; ICU, intensive care unit; BMI, body mass index; SES, social economics status; NDI, 2010 Neighborhood Socioeconomic Disadvantage Index.

^a^ The population density is used only in the tested positive/susceptibility model as a covariate.

**eTable 2.** Odds Ratios of COVID-19 Outcomes From Logistic Regression for the Full Cohort

| **Positive (1) vs Comparison Group (0)** | | **Unadjusted**  **(n_0_=,29373 n_1_=2582)** | **Adjustment 1**  **(n_0_=24051, n_1_=2300)** | **Adjustment 2**  **(n_0_=24051, n_1_=2300)** | **Adjustment 3**  **(n_0_=22988, n_1_=1960)** |
| --- | --- | --- | --- | --- | --- |
| **Variable** | | **OR (95% CI)** |  |  |  |
| **Age (unit: 10-year)** | | 1.08 (1.06, 1.09) | 1.14 (1.12, 1.16) | 1.14 (1.12, 1.16) | 0.97 (0.94, 0.99) |
| **Age Range**  REF: [18,35) | [0,18) | 0.25 (0.21, 0.31) | 0.24 (0.20, 0.30) | 0.25 (0.20, 0.30) | 0.31 (0.24, 0.38) |
|  | [35,50) | 1.33 (1.18, 1.50) | 1.51 (1.32, 1.72) | 1.52 (1.33, 1.73) | 1.14 (0.99, 1.32) |
|  | [50,65) | 1.27 (1.13, 1.42) | 1.56 (1.38, 1.77) | 1.57 (1.38, 1.78) | 0.85 (0.73, 0.98) |
|  | [65,80) | 0.89 (0.79, 1.01) | 1.22 (1.06, 1.40) | 1.22 (1.06, 1.41) | 0.53 (0.44, 0.62) |
|  | [80,100) | 0.75 (0.62, 0.91) | 1.04 (0.84, 1.29) | 1.04 (0.84, 1.29) | 0.39 (0.31, 0.50) |
| **Male Sex** | | 0.95 (0.88, 1.03) | 0.93 (0.85, 1.02) | 0.93 (0.85, 1.02) | 0.87 (0.79, 0.96) |
| **BMI** | | 1.04 (1.03, 1.04) | 1.03 (1.02, 1.04) | 1.03 (1.03, 1.04) | 1.02 (1.01, 1.02) |
| **BMI Range**  REF: [18.5,25) | <18.5 | 0.66 (0.43, 1.03) | 0.70 (0.45, 1.09) | 0.71 (0.46, 1.12) | 0.71 (0.44, 1.14) |
|  | [25,30) | 1.36 (1.20, 1.54) | 1.36 (1.19, 1.55) | 1.36 (1.19, 1.56) | 1.31 (1.14, 1.52) |
|  | >=30 | 1.95 (1.74, 2.19) | 1.82 (1.61, 2.07) | 1.85 (1.63, 2.10) | 1.50 (1.31, 1.72) |
| **Ever-Smoker** | | 1.01 (0.92, 1.11) | 0.83 (0.74, 0.92) | 0.85 (0.76, 0.94) | 0.80 (0.71, 0.90) |
| **Smoking Status**  REF: Never-Smoker | Past-Smoker | 1.26 (1.14, 1.40) | 1.05 (0.93, 1.18) | 1.06 (0.94, 1.19) | 0.95 (0.84, 1.08) |
|  | Current-Smoker | 0.46 (0.38, 0.57) | 0.40 (0.32, 0.49) | 0.41 (0.33, 0.51) | 0.45 (0.35, 0.57) |
| **Alcohol Consumption** | | 1.60 (1.44, 1.78) | 1.65 (1.47, 1.84) | 1.62 (1.45, 1.81) | 1.69 (1.50, 1.90) |
| **Race/Ethnicity**  REF: White | Black | 3.92 (3.53, 4.35) | 3.74 (3.35, 4.19) | 4.02 (3.56, 4.54) | 3.21 (2.80, 3.67) |
|  | Other / Known Ethnicity | 1.22 (1.06, 1.40) | 1.30 (1.12, 1.52) | 1.31 (1.12, 1.52) | 1.25 (1.06, 1.48) |
|  | Other / Unknown Ethnicity | 0.60 (0.53, 0.69) | 0.50 (0.43, 0.58) | 0.51 (0.43, 0.59) | 0.61 (0.50, 0.74) |
| **SES** | Population density (1000-people/mi^2^) | 7.54 (4.63, 12.3) | 0.40 (0.22, 0.73) | 0.40 (0.22, 0.73) | 0.192 (0.0975, 0.379) |
|  | NDI | 1.09 (1.07, 1.11) | 1.07 (1.05, 1.08) | 1.08 (1.06, 1.09) | 1.07 (1.05, 1.10) |
| **Comorbidity Score** | | 1.69 (1.64, 1.74) | 1.70 (1.64, 1.76) | 1.71 (1.64, 1.77) | NA |
| **Comorbidities** | Respiratory | 4.10 (3.71, 4.54) | 3.76 (3.37, 4.20) | 3.74 (3.34, 4.17) | NA |
|  | Circulatory | 3.29 (2.99, 3.61) | 3.07 (2.75, 3.43) | 3.09 (2.77, 3.45) | NA |
|  | Any Cancer | 1.30 (1.17, 1.44) | 1.19 (1.05, 1.35) | 1.19 (1.05, 1.34) | NA |
|  | Type 2 Diabetes | 2.86 (2.55, 3.21) | 2.21 (1.94, 2.52) | 2.26 (1.98, 2.57) | NA |
|  | Kidney | 3.92 (3.44, 4.45) | 2.90 (2.51, 3.35) | 2.96 (2.56, 3.43) | NA |
|  | Liver | 3.52 (2.96, 4.18) | 3.01 (2.50, 3.63) | 3.04 (2.52, 3.67) | NA |
|  | Autoimmune | 2.74 (2.43, 3.09) | 2.33 (2.05, 2.65) | 2.33 (2.05, 2.65) | NA |
| **Hospitalization (1) vs Not (0)** | | **Unadjusted**  **(n_0_=1852, n_1_=719)** | **Adjustment 1**  **(n_0_=1852, n_1_=719)** | **Adjustment 2**  **(n_0_=1628, n_1_=662)** | **Adjustment 3**  **(n_0_=1479, n_1_=471)** |
| **Age (unit: 10-year)** | | 1.56 (1.48, 1.64) | 1.58 (1.5, 1.67) | 1.55 (1.46, 1.64) | 1.46 (1.36, 1.56) |
| **Age Range**  REF: [18,35) | [0,18) | 1.11 (0.61, 2.04) | 1.09 (0.589, 2) | 1.07 (0.562, 2.04) | 1.24 (0.604, 2.56) |
|  | [35,50) | 2.42 (1.75, 3.35) | 2.25 (1.62, 3.12) | 1.92 (1.36, 2.71) | 1.51 (1, 2.28) |
|  | [50,65) | 4.33 (3.20, 5.86) | 4.19 (3.08, 5.7) | 3.72 (2.69, 5.14) | 2.62 (1.78, 3.85) |
|  | [65,80) | 8.35 (6.07, 11.5) | 8.39 (6.05, 11.6) | 7.17 (5.08, 10.1) | 4.62 (3.07, 6.95) |
|  | [80,100) | 14.5 (9.38, 22.5) | 16.4 (10.4, 25.7) | 14.4 (8.89, 23.4) | 13.3 (7.51, 23.4) |
| **Male Sex** | | 1.84 (1.55, 2.19) | 1.92 (1.59, 2.33) | 2.02 (1.65, 2.47) | 1.84 (1.46, 2.31) |
| **BMI** | | 1.04 (1.03, 1.05) | 1.03 (1.02, 1.05) | 1.03 (1.02, 1.05) | 1.02 (1, 1.04) |
| **BMI Range**  REF: [18.5,25) | <18.5 | 2.72 (1.13, 6.54) | 3.03 (1.05, 8.79) | 2.86 (0.971, 8.42) | 3.61 (1.17, 11.2) |
|  | [25,30) | 1.89 (1.44, 2.48) | 1.23 (0.905, 1.67) | 1.32 (0.952, 1.82) | 1.47 (1.02, 2.14) |
|  | >=30 | 2.16 (1.68, 2.78) | 1.62 (1.22, 2.16) | 1.64 (1.21, 2.22) | 1.6 (1.12, 2.27) |
| **Ever-Smoker** | | 1.71 (1.41, 2.08) | 1.12 (0.901, 1.4) | 1.05 (0.838, 1.33) | 1.09 (0.848, 1.4) |
| **Smoking Status**  REF: Never-Smoker | Past-Smoker | 1.87 (1.52, 2.29) | 1.16 (0.917, 1.46) | 1.13 (0.889, 1.44) | 1.15 (0.883, 1.49) |
|  | Current-Smoker | 0.99 (0.62, 1.58) | 0.949 (0.577, 1.56) | 0.678 (0.388, 1.19) | 0.781 (0.427, 1.43) |
| **Alcohol Consumption** | | 0.84 (0.66, 1.05) | 0.839 (0.652, 1.08) | 0.853 (0.658, 1.11) | 0.771 (0.587, 1.01) |
| **Race/Ethnicity**  REF: White | Black | 2.32 (1.90, 2.83) | 2.63 (2.11, 3.28) | 2.1 (1.63, 2.7) | 1.85 (1.39, 2.47) |
|  | Other / Known Ethnicity | 1.11 (0.81, 1.51) | 1.42 (1.02, 1.99) | 1.31 (0.923, 1.87) | 1.15 (0.751, 1.75) |
|  | Other / Unknown Ethnicity | 1.06 (0.78, 1.44) | 0.911 (0.656, 1.27) | 1.02 (0.705, 1.48) | 1.11 (0.686, 1.78) |
| **SES** | Population density (1000-people/mi^2^) | 64.8 (25, 168) | 21.6 (6.94, 67.5) | 21.6 (6.94, 67.5) | 11.7 (3.08, 44.4) |
|  | NDI | 1.15 (1.11, 1.19) | 1.11 (1.07, 1.15) | 1.07 (1.03, 1.12) | 1.07 (1.02, 1.13) |
| **Comorbidity Score** | | 1.56 (1.46, 1.67) | 1.26 (1.17, 1.36) | 1.24 (1.15, 1.35) | NA |
| **Comorbidities** | Respiratory | 1.32 (1.03, 1.68) | 1.11 (0.847, 1.45) | 1.14 (0.863, 1.52) | NA |
|  | Circulatory | 3.26 (2.49, 4.25) | 1.66 (1.24, 2.23) | 1.58 (1.16, 2.15) | NA |
|  | Any Cancer | 2 (1.6, 2.5) | 1.18 (0.912, 1.51) | 1.17 (0.902, 1.53) | NA |
|  | Type 2 Diabetes | 3.58 (2.85, 4.49) | 1.8 (1.4, 2.31) | 1.79 (1.39, 2.32) | NA |
|  | Kidney | 6.62 (5.16, 8.5) | 3.44 (2.62, 4.52) | 3.2 (2.41, 4.24) | NA |
|  | Liver | 1.79 (1.28, 2.5) | 1.33 (0.925, 1.9) | 1.24 (0.856, 1.8) | NA |
|  | Autoimmune | 1.4 (1.09, 1.8) | 1.1 (0.838, 1.45) | 1.06 (0.803, 1.41) | NA |
| **ICU (1) vs Not (0)** | | **Unadjusted**  **(n_0_=2194, n_1_=377)** | **Adjustment 1**  **(n_0_=2194, n_1_=377)** | **Adjustment 2**  **(n_0_=1941, n_1_=349)** | **Adjustment 3**  **(n_0_=1711, n_1_=239)** |
| **Age (unit: 10-year)** | | 1.44 (1.35, 1.53) | 1.43 (1.34, 1.52) | 1.41 (1.32, 1.51) | 1.34 (1.23, 1.47) |
| **Age Range**  REF: [18,35) | [0,18) | 1.61 (0.78, 3.31) | 1.57 (0.759, 3.26) | 1.46 (0.677, 3.15) | 2.21 (0.906, 5.38) |
|  | [35,50) | 1.95 (1.25, 3.04) | 1.84 (1.17, 2.87) | 1.45 (0.912, 2.32) | 1.42 (0.778, 2.58) |
|  | [50,65) | 3.9 (2.61, 5.82) | 3.65 (2.44, 5.47) | 3.03 (2, 4.6) | 2.56 (1.48, 4.44) |
|  | [65,80) | 7.38 (4.92, 11.1) | 6.78 (4.5, 10.2) | 5.8 (3.79, 8.87) | 4.39 (2.51, 7.7) |
|  | [80,100) | 6.36 (3.77, 10.7) | 6.22 (3.66, 10.6) | 5.23 (2.98, 9.2) | 5.39 (2.67, 10.9) |
| **Male Sex** | | 2.21 (1.76, 2.76) | 2.24 (1.78, 2.83) | 2.41 (1.88, 3.08) | 2.33 (1.74, 3.13) |
| **BMI** | | 1.03 (1.02, 1.05) | 1.03 (1.02, 1.05) | 1.03 (1.01, 1.05) | 1.01 (0.99, 1.03) |
| **BMI Range**  REF: [18.5,25) | <18.5 | 1.73 (0.584, 5.13) | 1.48 (0.417, 5.22) | 1.25 (0.336, 4.67) | 2.96 (0.755, 11.6) |
|  | [25,30) | 1.47 (1.05, 2.06) | 0.964 (0.67, 1.39) | 1.04 (0.705, 1.53) | 1.17 (0.734, 1.86) |
|  | >=30 | 1.61 (1.18, 2.2) | 1.23 (0.875, 1.72) | 1.23 (0.858, 1.77) | 1.08 (0.688, 1.68) |
| **Ever-Smoker** | | 2.03 (1.58, 2.61) | 1.35 (1.03, 1.77) | 1.24 (0.94, 1.65) | 1.29 (0.941, 1.77) |
| **Smoking Status**  REF: Never-Smoker | Past-Smoker | 2.2 (1.7, 2.85) | 1.4 (1.06, 1.86) | 1.32 (0.988, 1.77) | 1.35 (0.968, 1.87) |
|  | Current-Smoker | 1.19 (0.642, 2.21) | 1.09 (0.576, 2.05) | 0.817 (0.398, 1.68) | 0.997 (0.46, 2.16) |
| **Alcohol Consumption** | | 0.878 (0.646, 1.19) | 0.823 (0.595, 1.14) | 0.838 (0.6, 1.17) | 0.813 (0.573, 1.15) |
| **Race/Ethnicity**  REF: White | Black | 1.97 (1.54, 2.53) | 2.1 (1.62, 2.73) | 1.59 (1.18, 2.15) | 1.33 (0.931, 1.9) |
|  | Other / Known Ethnicity | 0.665 (0.415, 1.06) | 0.782 (0.481, 1.27) | 0.743 (0.452, 1.22) | 0.818 (0.454, 1.48) |
|  | Other / Unknown Ethnicity | 1.45 (1.02, 2.07) | 1.35 (0.934, 1.96) | 1.36 (0.897, 2.07) | 1.43 (0.809, 2.52) |
| **SES** | Population density (1000-people/mi^2^) | 62.6 (20.8, 188) | 30.7 (8.38, 112) | 30.7 (8.38, 112) | 20.7 (4.17, 102) |
|  | NDI | 1.15 (1.1, 1.19) | 1.12 (1.07, 1.17) | 1.07 (1.02, 1.13) | 1.1 (1.03, 1.17) |
| **Comorbidity Score** | | 1.6 (1.47, 1.75) | 1.36 (1.23, 1.5) | 1.34 (1.21, 1.48) | NA |
| **Comorbidities** | Respiratory | 1.46 (1.05, 2.04) | 1.3 (0.914, 1.85) | 1.36 (0.942, 1.97) | NA |
|  | Circulatory | 3.24 (2.22, 4.72) | 1.77 (1.18, 2.65) | 1.62 (1.07, 2.45) | NA |
|  | Any Cancer | 2.03 (1.53, 2.69) | 1.27 (0.928, 1.73) | 1.32 (0.959, 1.83) | NA |
|  | Type 2 Diabetes | 3.58 (2.71, 4.72) | 1.96 (1.45, 2.65) | 1.89 (1.39, 2.59) | NA |
|  | Kidney | 6.9 (5.19, 9.18) | 3.85 (2.82, 5.27) | 3.64 (2.64, 5.02) | NA |
|  | Liver | 1.87 (1.24, 2.81) | 1.44 (0.94, 2.22) | 1.34 (0.861, 2.09) | NA |
|  | Autoimmune | 1.58 (1.16, 2.16) | 1.39 (0.995, 1.93) | 1.3 (0.927, 1.84) | NA |
| **Deceased (1) vs Alive (0)** | | **Unadjusted**  **(n_0_=2453, n_1_=129)** | **Adjustment 1**  **(n_0_=2453, n_1_=129)** | **Adjustment 2**  **(n_0_=2186, n_1_=114)** | **Adjustment 3**  **(n_0_=1876, n_1_=84)** |
| **Age (unit: 10-year)** | | 1.44 (1.35, 1.53) | 1.41 (1.32, 1.51) | 2.0 (1.75, 2.30) | 2.1 (1.74, 2.52) |
| **Age Range**  REF: [18,35) | [0,18) | 1.61 (0.78, 3.31) | 1.46 (0.68, 3.15) | 0.67 (0.03, 12.8) | 1.09 (0.05, 22.1) |
|  | [35,50) | 1.95 (1.25, 3.04) | 1.45 (0.912, 2.32) | 2.65 (0.791, 8.87) | 1.74 (0.41, 7.38) |
|  | [50,65) | 3.90 (2.61, 5.82) | 3.03 (2.00, 4.60) | 6.55 (2.17, 19.8) | 2.95 (0.772, 11.3) |
|  | [65,80) | 7.38 (4.92, 11.1) | 5.8 (3.79, 8.87) | 14.5 (4.85, 43.1) | 7.14 (1.91, 26.7) |
|  | [80,100) | 6.36 (3.77, 10.7) | 5.23 (2.98, 9.2) | 50.7 (16.4, 156) | 36.5 (9.49, 140) |
| **Male Sex** | | 2.21 (1.76, 2.76) | 2.41 (1.88, 3.08) | 2.28 (1.51, 3.43) | 1.93 (1.19, 3.13) |
| **BMI** | | 1.03 (1.02, 1.05) | 1.03 (1.01, 1.05) | 1.02 (0.996, 1.04) | 1.02 (0.985, 1.05) |
| **BMI Range**  REF: [18.5,25) | <18.5 | 1.73 (0.58, 5.13) | 1.25 (0.336, 4.67) | 0.802 (0.0939, 6.85) | 6.34 (0.652, 61.6) |
|  | [25,30) | 1.47 (1.05, 2.06) | 1.04 (0.705, 1.53) | 1.41 (0.72, 2.77) | 1.54 (0.655, 3.62) |
|  | >=30 | 1.61 (1.18, 2.20) | 1.23 (0.858, 1.77) | 1.97 (1.03, 3.76) | 2.08 (0.906, 4.79) |
| **Ever-Smoker** | | 2.03 (1.58, 2.61) | 1.24 (0.94, 1.65) | 2.06 (1.21, 3.5) | 1.45 (0.82, 2.58) |
| **Smoking Status**  REF: Never-Smoker | Past-Smoker | 2.20 (1.70, 2.85) | 1.32 (0.988, 1.77) | 2.08 (1.21, 3.56) | 1.5 (0.843, 2.68) |
|  | Current-Smoker | 1.19 (0.64, 2.21) | 0.817 (0.398, 1.68) | 2.26 (0.57, 8.94) | 1.09 (0.182, 6.46) |
| **Alcohol Consumption** | | 0.88 (0.65, 1.19) | 0.838 (0.6, 1.17) | 0.899 (0.492, 1.64) | 1.05 (0.562, 1.95) |
| **Race/Ethnicity**  REF: White | Black | 1.97 (1.54, 2.53) | 1.59 (1.18, 2.15) | 1.43 (0.871, 2.36) | 1.46 (0.813, 2.64) |
|  | Other / Known Ethnicity | 0.67 (0.42, 1.06) | 0.743 (0.452, 1.22) | 0.834 (0.348, 2) | 1.26 (0.479, 3.32) |
|  | Other / Unknown Ethnicity | 1.45 (1.02, 2.07) | 1.36 (0.897, 2.07) | 2 (1.11, 3.63) | 5.44 (2.63, 11.3) |
| **SES** | Population density (1000-people/mi^2^) | 62.6 (20.8, 188) | 30.7 (8.38, 112) | 38.6 (5.16, 288) | 110 (9.92, 1230) |
|  | NDI | 1.15 (1.10, 1.19) | 1.07 (1.02, 1.13) | 1.09 (0.999, 1.18) | 1.11 (1.01, 1.23) |
| **Comorbidity Score** | | 1.60 (1.47, 1.75) | 1.34 (1.21, 1.48) | 1.52 (1.29, 1.79) | NA |
| **Comorbidities** | Respiratory | 1.46 (1.05, 2.04) | 1.36 (0.94, 1.97) | 2.50 (1.22, 5.13) | NA |
|  | Circulatory | 3.24 (2.22, 4.72) | 1.62 (1.07, 2.45) | 2.91 (1.09, 7.78) | NA |
|  | Any Cancer | 2.03 (1.53, 2.69) | 1.32 (0.96, 1.83) | 1.62 (0.99, 2.65) | NA |
|  | Type 2 Diabetes | 3.58 (2.71, 4.72) | 1.89 (1.39, 2.59) | 3.39 (2.07, 5.57) | NA |
|  | Kidney | 6.90 (5.19, 9.18) | 3.64 (2.64, 5.02) | 3.82 (2.31, 6.31) | NA |
|  | Liver | 1.87 (1.24, 2.81) | 1.34 (0.86, 2.09) | 1.67 (0.85, 3.26) | NA |
|  | Autoimmune | 1.58 (1.16, 2.16) | 1.30 (0.93, 1.84) | 0.93 (0.52, 1.67) | NA |

Abbreviations: OR, odds ratio; ICU, intensive care unit; BMI, body mass index; NA, not applicable; REF, reference group; SES, social economics status; NDI, 2010 Neighborhood Socioeconomic Disadvantage Index; adjustment 0, unadjusted; adjustment 1, age+sex+race/ethnicity+(persons per mile^2^ in susceptibility model only); adjustment 2, adjustment 1+NDI; adjustment 3, adjustment 2+comorbidity score.

The model used was: $logit P\left( Y_{\mathrm{COVID}}=1|X, adjustment \right)=\beta_{0}+\beta_{X}X+\beta_{\mathrm{adjust}}\mathrm{adjustment}_{j}$. Here $Y_{\mathrm{COVID}}$ is various COVID-19 related outcomes under consideration (i.e., COVID-19 positive, hospitalization and ICU admission); $X$ is the variable/risk factor of interest; and $\mathrm{adjustment}_{j}$, j = 0, …,3 are the four nested covariate adjustment models listed in eTable 1.

**eTable 3. Characteristics of the COVID-19 Tested or Diagnosed Cohort, Stratified by Quarters**

| 3A. First Quarter (March 10, 2020, to March 31, 2020) | | | | | | | |
| --- | --- | --- | --- | --- | --- | --- | --- |
|  |  |  | **Positive Results** | | | |  |
|  | **Overall** | **Negative Results** | **Overall** | **Hospitalized** | **ICU** | **Deceased** | **Comparison Group** |
| Variable | (n = 2258) | (n = 1760) | (n = 498) | (n = 209) | (n = 123) | (n = 39) | (n = 29383) |
| Age, y |  |  |  |  |  |  |  |
| Mean (SD) | 44.5 (19.9) | 42.8 (20.3) | 50.2 (17.1) | 59.9 (15.8) | 62.8 (14.6) | 69.3 (12.7) | 43.2 (24.4) |
| Median (IQR) | 44 (28) | 42 (28) | 51 (27) | 61 (20) | 64 (16) | 70 (19.5) | 43 (42) |
| <18 | 166 (7.4) | 163 (9.3) | 3 (0.6) | 1 (0.5) | 1 (0.8) | 0 (0) | 5345 (18.2) |
| [18,35) | 569 (25.2) | 467 (26.5) | 102 (20.5) | 13 (6.2) | 3 (2.4) | 1 (2.6) | 6731 (22.9) |
| [35,50) | 601 (26.6) | 474 (26.9) | 127 (25.5) | 36 (17.2) | 15 (12.2) | 1 (2.6) | 4593 (15.6) |
| [50,65) | 551 (24.4) | 396 (22.5) | 155 (31.1) | 74 (35.4) | 44 (35.8) | 10 (25.6) | 5659 (19.3) |
| [65,80) | 283 (12.5) | 199 (11.3) | 84 (16.9) | 59 (28.2) | 43 (35) | 15 (38.5) | 5128 (17.5) |
| >=80 | 88 (3.9) | 61 (3.5) | 27 (5.4) | 26 (12.4) | 17 (13.8) | 12 (30.8) | 1916 (6.5) |
| Gender | 859 (38) | 635 (36.1) | 224 (45) | 118 (56.5) | 81 (65.9) | 26 (66.7) | 13581 (46.3) |
| Primary Care in MM | 1396 (61.8) | 1118 (63.5) | 278 (55.8) | 81 (38.8) | 46 (37.4) | 15 (38.5) | 4318 (14.7) |
| BMI |  |  |  |  |  |  |  |
| Mean (SD) | 29.9 (8.2) | 29.2 (7.4) | 32 (10.1) | 33.3 (12.5) | 33.8 (15.1) | 30.2 (6.5) | 28.5 (7.3) |
| <18.5 | 29 (1.5) | 26 (1.7) | 3 (0.6) | 2 (1) | 1 (0.8) | 1 (2.6) | 357 (2.2) |
| [18.5,25) | 553 (27.9) | 464 (30.6) | 89 (19.1) | 27 (13.4) | 19 (15.8) | 6 (15.8) | 5278 (32.8) |
| [25,30) | 561 (28.3) | 429 (28.3) | 132 (28.3) | 60 (29.9) | 37 (30.8) | 13 (34.2) | 5020 (31.2) |
| >=30 | 841 (42.4) | 599 (39.5) | 242 (51.9) | 112 (55.7) | 63 (52.5) | 18 (47.4) | 5455 (33.9) |
| Smoking Status |  |  |  |  |  |  |  |
| Never | 1359 (65.1) | 1051 (63.8) | 308 (69.7) | 112 (60.2) | 52 (50.5) | 11 (40.7) | 15002 (68.9) |
| Past | 577 (27.6) | 456 (27.7) | 121 (27.4) | 70 (37.6) | 49 (47.6) | 16 (59.3) | 4615 (21.2) |
| Current | 153 (7.3) | 140 (8.5) | 13 (2.9) | 4 (2.2) | 2 (1.9) | 0 (0) | 2168 (10) |
| Ever | 730 (34.9) | 596 (36.2) | 134 (30.3) | 74 (39.8) | 51 (49.5) | 16 (59.3) | 6783 (31.1) |
| Alcohol consumption | 1199 (69.5) | 951 (69.4) | 248 (70.1) | 92 (68.7) | 53 (73.6) | 15 (68.2) | 8542 (55) |
| Race/ethnicity |  |  |  |  |  |  |  |
| White | 1492 (66.1) | 1284 (73) | 208 (41.8) | 75 (35.9) | 48 (39) | 15 (38.5) | 18529 (63.1) |
| Black | 433 (19.2) | 222 (12.6) | 211 (42.4) | 106 (50.7) | 60 (48.8) | 22 (56.4) | 2165 (7.4) |
| Other^b^ | 238 (10.5) | 183 (10.4) | 55 (11) | 17 (8.1) | 7 (5.7) | 1 (2.6) | 2731 (9.3) |
| Unknown^c^ | 95 (4.2) | 71 (4) | 24 (4.8) | 11 (5.3) | 8 (6.5) | 1 (2.6) | 5958 (20.3) |
| NDI, mean (SD) | 0.11 (0.08) | 0.1 (0.07) | 0.14 (0.1) | 0.16 (0.11) | 0.16 (0.11) | 0.18 (0.09) | 0.11 (0.08) |
| Population density, persons/square mile | 2680.1 (2456.7) | 2493.6 (2446.5) | 3336.1 (2381.3) | 3846.1 (2521.5) | 3865.4 (2523.2) | 4758.3 (2384.7) | 2335.1 (2495.1) |
| Comorbidity score, mean (SD) | 2.4 (1.6) | 2.5 (1.6) | 2.4 (1.6) | 3.1 (1.7) | 3.2 (1.7) | 4.2 (1.6) | 1.3 (1.2) |

| 3B. Second Quarter (April 1, 2020, to June 30, 2020) | | | | | | | |
| --- | --- | --- | --- | --- | --- | --- | --- |
|  |  |  | **Positive Results** | | | |  |
|  | **Overall** | **Negative Results** | **Overall** | **Hospitalized** | **ICU** | **Deceased** | **Comparison Group** |
| Variable | (n = 2258) | (n = 1760) | (n = 498) | (n = 209) | (n = 123) | (n = 39) | (n = 29383) |
| Age, y |  |  |  |  |  |  |  |
| Mean (SD) | 47.3 (23.3) | 47.1 (23.3) | 51.8 (20.4) | 57.2 (19.2) | 56.1 (19) | 69.5 (15.4) | 43.2 (24.4) |
| Median (IQR) | 51 (36) | 50 (36) | 54 (31) | 59 (25) | 59 (28) | 73 (23.5) | 43 (42) |
| <18 | 2866 (12.2) | 2832 (12.6) | 34 (3.9) | 12 (3.2) | 8 (3.9) | 0 (0) | 5345 (18.2) |
| [18,35) | 4536 (19.4) | 4372 (19.4) | 164 (18.6) | 41 (11) | 28 (13.7) | 2 (3) | 6731 (22.9) |
| [35,50) | 3966 (16.9) | 3793 (16.8) | 173 (19.7) | 65 (17.4) | 29 (14.1) | 6 (9) | 4593 (15.6) |
| [50,65) | 5526 (23.6) | 5283 (23.4) | 243 (27.6) | 108 (28.9) | 60 (29.3) | 16 (23.9) | 5659 (19.3) |
| [65,80) | 5095 (21.7) | 4899 (21.7) | 196 (22.3) | 106 (28.3) | 66 (32.2) | 21 (31.3) | 5128 (17.5) |
| >=80 | 1441 (6.2) | 1371 (6.1) | 70 (8) | 42 (11.2) | 14 (6.8) | 22 (32.8) | 1916 (6.5) |
| Gender | 10420 (44.5) | 10005 (44.4) | 415 (47.2) | 213 (57) | 121 (59) | 44 (65.7) | 13581 (46.3) |
| Primary Care in MM | 13597 (58) | 13152 (58.3) | 445 (50.6) | 139 (37.2) | 73 (35.6) | 19 (28.4) | 4318 (14.7) |
| BMI |  |  |  |  |  |  |  |
| Mean (SD) | 29.5 (7.8) | 29.4 (7.7) | 31.3 (8.4) | 32.3 (8.9) | 32.6 (9.1) | 31.7 (7) | 28.5 (7.3) |
| <18.5 | 381 (1.9) | 372 (2) | 9 (1.2) | 5 (1.4) | 2 (1) | 0 (0) | 357 (2.2) |
| [18.5,25) | 5420 (27.6) | 5261 (27.9) | 159 (20.8) | 59 (16.5) | 34 (17.4) | 10 (15.4) | 5278 (32.8) |
| [25,30) | 6123 (31.2) | 5899 (31.2) | 224 (29.3) | 107 (29.9) | 54 (27.7) | 19 (29.2) | 5020 (31.2) |
| >=30 | 7729 (39.3) | 7356 (38.9) | 373 (48.8) | 187 (52.2) | 105 (53.8) | 36 (55.4) | 5455 (33.9) |
| Smoking Status |  |  |  |  |  |  |  |
| Never | 13316 (60.2) | 12849 (60.1) | 467 (64.2) | 196 (61.6) | 89 (57.1) | 18 (40.9) | 15002 (68.9) |
| Past | 6684 (30.2) | 6469 (30.3) | 215 (29.6) | 106 (33.3) | 58 (37.2) | 23 (52.3) | 4615 (21.2) |
| Current | 2107 (9.5) | 2062 (9.6) | 45 (6.2) | 16 (5) | 9 (5.8) | 3 (6.8) | 2168 (10) |
| Ever | 8791 (39.8) | 8531 (39.9) | 260 (35.8) | 122 (38.4) | 67 (42.9) | 26 (59.1) | 6783 (31.1) |
| Alcohol consumption | 11578 (67.7) | 11258 (68) | 320 (60) | 118 (57) | 65 (58) | 18 (58.1) | 8542 (55) |
| Race/ethnicity |  |  |  |  |  |  |  |
| White | 17213 (73.5) | 16775 (74.4) | 438 (49.8) | 179 (47.9) | 102 (49.8) | 35 (52.2) | 18529 (63.1) |
| Black | 2825 (12.1) | 2575 (11.4) | 250 (28.4) | 124 (33.2) | 66 (32.2) | 14 (20.9) | 2165 (7.4) |
| Other^b^ | 1997 (8.5) | 1908 (8.5) | 89 (10.1) | 36 (9.6) | 11 (5.4) | 5 (7.5) | 2731 (9.3) |
| Unknown^c^ | 1395 (6) | 1292 (5.7) | 103 (11.7) | 35 (9.4) | 26 (12.7) | 13 (19.4) | 5958 (20.3) |
| NDI, mean (SD) | 0.1 (0.08) | 0.1 (0.08) | 0.13 (0.1) | 0.14 (0.1) | 0.15 (0.11) | 0.14 (0.1) | 0.11 (0.08) |
| Population density, persons/square mile | 2374.9 (2389.3) | 2338.6 (2372.9) | 3284.7 (2611.6) | 3577.7 (2706.6) | 3674 (2749.3) | 3714.9 (2894) | 2335.1 (2495.1) |
| Comorbidity score, mean (SD) | 2.4 (1.6) | 2.4 (1.6) | 2.6 (1.6) | 3.2 (1.7) | 3.4 (1.6) | 3.8 (1.5) | 1.3 (1.2) |

| 3C. Third Quarter (July 1, 2020, to September 2, 2020) | | | | | | | |
| --- | --- | --- | --- | --- | --- | --- | --- |
|  |  |  | **Positive Results** | | | |  |
|  | **Overall** | **Negative Results** | **Overall** | **Hospitalized** | **ICU** | **Deceased** | **Comparison Group** |
| Variable | (n = 2258) | (n = 1760) | (n = 498) | (n = 209) | (n = 123) | (n = 39) | (n = 29383) |
| Age, y |  |  |  |  |  |  |  |
| Mean (SD) | 47.3 (23.3) | 47.1 (23.3) | 51.8 (20.4) | 57.2 (19.2) | 56.1 (19) | 69.5 (15.4) | 43.2 (24.4) |
| Median (IQR) | 51 (36) | 50 (36) | 54 (31) | 59 (25) | 59 (28) | 73 (23.5) | 43 (42) |
| <18 | 2866 (12.2) | 2832 (12.6) | 34 (3.9) | 12 (3.2) | 8 (3.9) | 0 (0) | 5345 (18.2) |
| [18,35) | 4536 (19.4) | 4372 (19.4) | 164 (18.6) | 41 (11) | 28 (13.7) | 2 (3) | 6731 (22.9) |
| [35,50) | 3966 (16.9) | 3793 (16.8) | 173 (19.7) | 65 (17.4) | 29 (14.1) | 6 (9) | 4593 (15.6) |
| [50,65) | 5526 (23.6) | 5283 (23.4) | 243 (27.6) | 108 (28.9) | 60 (29.3) | 16 (23.9) | 5659 (19.3) |
| [65,80) | 5095 (21.7) | 4899 (21.7) | 196 (22.3) | 106 (28.3) | 66 (32.2) | 21 (31.3) | 5128 (17.5) |
| >=80 | 1441 (6.2) | 1371 (6.1) | 70 (8) | 42 (11.2) | 14 (6.8) | 22 (32.8) | 1916 (6.5) |
| Gender | 10420 (44.5) | 10005 (44.4) | 415 (47.2) | 213 (57) | 121 (59) | 44 (65.7) | 13581 (46.3) |
| Primary Care in MM | 13597 (58) | 13152 (58.3) | 445 (50.6) | 139 (37.2) | 73 (35.6) | 19 (28.4) | 4318 (14.7) |
| BMI |  |  |  |  |  |  |  |
| Mean (SD) | 29.5 (7.8) | 29.4 (7.7) | 31.3 (8.4) | 32.3 (8.9) | 32.6 (9.1) | 31.7 (7) | 28.5 (7.3) |
| <18.5 | 381 (1.9) | 372 (2) | 9 (1.2) | 5 (1.4) | 2 (1) | 0 (0) | 357 (2.2) |
| [18.5,25) | 5420 (27.6) | 5261 (27.9) | 159 (20.8) | 59 (16.5) | 34 (17.4) | 10 (15.4) | 5278 (32.8) |
| [25,30) | 6123 (31.2) | 5899 (31.2) | 224 (29.3) | 107 (29.9) | 54 (27.7) | 19 (29.2) | 5020 (31.2) |
| >=30 | 7729 (39.3) | 7356 (38.9) | 373 (48.8) | 187 (52.2) | 105 (53.8) | 36 (55.4) | 5455 (33.9) |
| Smoking Status |  |  |  |  |  |  |  |
| Never | 13316 (60.2) | 12849 (60.1) | 467 (64.2) | 196 (61.6) | 89 (57.1) | 18 (40.9) | 15002 (68.9) |
| Past | 6684 (30.2) | 6469 (30.3) | 215 (29.6) | 106 (33.3) | 58 (37.2) | 23 (52.3) | 4615 (21.2) |
| Current | 2107 (9.5) | 2062 (9.6) | 45 (6.2) | 16 (5) | 9 (5.8) | 3 (6.8) | 2168 (10) |
| Ever | 8791 (39.8) | 8531 (39.9) | 260 (35.8) | 122 (38.4) | 67 (42.9) | 26 (59.1) | 6783 (31.1) |
| Alcohol consumption | 11578 (67.7) | 11258 (68) | 320 (60) | 118 (57) | 65 (58) | 18 (58.1) | 8542 (55) |
| Race/ethnicity |  |  |  |  |  |  |  |
| White | 17213 (73.5) | 16775 (74.4) | 438 (49.8) | 179 (47.9) | 102 (49.8) | 35 (52.2) | 18529 (63.1) |
| Black | 2825 (12.1) | 2575 (11.4) | 250 (28.4) | 124 (33.2) | 66 (32.2) | 14 (20.9) | 2165 (7.4) |
| Other^b^ | 1997 (8.5) | 1908 (8.5) | 89 (10.1) | 36 (9.6) | 11 (5.4) | 5 (7.5) | 2731 (9.3) |
| Unknown^c^ | 1395 (6) | 1292 (5.7) | 103 (11.7) | 35 (9.4) | 26 (12.7) | 13 (19.4) | 5958 (20.3) |
| NDI, mean (SD) | 0.1 (0.08) | 0.1 (0.08) | 0.13 (0.1) | 0.14 (0.1) | 0.15 (0.11) | 0.14 (0.1) | 0.11 (0.08) |
| Population density, persons/square mile | 2374.9 (2389.3) | 2338.6 (2372.9) | 3284.7 (2611.6) | 3577.7 (2706.6) | 3674 (2749.3) | 3714.9 (2894) | 2335.1 (2495.1) |
| Comorbidity score, mean (SD) | 2.4 (1.6) | 2.4 (1.6) | 2.6 (1.6) | 3.2 (1.7) | 3.4 (1.6) | 3.8 (1.5) | 1.3 (1.2) |
| Abbreviations: BMI, body mass index (calculated as weight in kilograms divided by height in meters squared); COVID-19, coronavirus disease 2019; ICU, intensive care unit; IQR, interquartile range; NDI, 2010 Neighborhood Socioeconomic Disadvantage Index; MM, Michigan Medicine.  a Percentages are reported as fraction of column totals excluding missing entries.  b Includes White Hispanic or unknown; Black Hispanic or unknown; Asian Hispanic,non-Hispanic, or unknown; Native American Hispanic, non-Hispanic, or unknown;Pacific Islander Hispanic, non-Hispanic, or unknown; and other Hispanic, non-Hispanic, or unknown.  c Includes missing race and/or ethnicity. | | | | | | | |

**eTable 4.** Descriptive Characteristics of the COVID-19 Tested or Diagnosed Cohort Stratified by White and Black Patients

| **White Patients** | | **Total Tested for COVID-19** | | | | | | **Matched Comparison Group**  **(n=18529)** |
| --- | --- | --- | --- | --- | --- | --- | --- | --- |
|  |  | **Overall**  **(n=38977)** | **Negative**  **(n=37566)** | **Tested Positive** | | | |  |
|  |  |  |  | **Overall**  **(n=1411)** | **Hospitalized**  **(n=326)** | **ICU**  **(n=172)** | **Deceased**  **(n=56)** |  |
| **Variable** | | **n (%)** |  |  |  |  |  |  |
| **Age**  **(in years)** | mean (SD) | 46.1 (23.3) | 46.0 (23.4) | 47.0 (20.3) | 59.9 (18.1) | 59.4 (18.5) | 72.2 (15.8) | 43.0 (24.5) |
|  | median [IQR] | 49.0 [37.0] | 49.0 [37.0] | 49.0 [34.0] | 63.0 [22.0] | 63.0 [22.0] | 77.0 [18.8] | 43.0 [42.0] |
| **Age Range** | [0,18) | 5020 (12.9) | 4951 (13.2) | 69 (4.9) | 7 (2.1) | 5 (2.9) | 0 (0) | 3450 (18.6) |
|  | [18,35) | 8304 (21.3) | 7929 (21.1) | 375 (26.6) | 31 (9.5) | 17 (9.9) | 3 (5.4) | 4232 (22.8) |
|  | [35,50) | 6345 (16.3) | 6058 (16.1) | 287 (20.3) | 40 (12.3) | 20 (11.6) | 3 (5.4) | 2816 (15.2) |
|  | [50,65) | 9063 (23.3) | 8702 (23.2) | 361 (25.6) | 100 (30.7) | 51 (29.7) | 8 (14.3) | 3560 (19.2) |
|  | [65,80) | 8177 (21.0) | 7928 (21.1) | 249 (17.6) | 105 (32.2) | 61 (35.5) | 18 (32.1) | 3320 (17.9) |
|  | [80,100) | 2068 (5.3) | 1998 (5.3) | 70 (5.0) | 43 (13.2) | 18 (10.5) | 24 (42.9) | 1151 (6.2) |
| **Male Sex** | | 17327 (44.5) | 16672 (44.4) | 655 (46.4) | 196 (60.1) | 114 (66.3) | 38 (67.9) | 8606 (46.4) |
| **Primary Care in MM** | | 23813 (61.1) | 22935 (61.1) | 878 (62.2) | 139 (42.6) | 81 (47.1) | 22 (39.3) | 2939 (15.9) |
| **BMI, mean (SD)** | |  |  |  |  |  |  |  |
| **BMI Range** | <18.5 | 29.0 (7.28) | 29.0 (7.27) | 29.6 (7.42) | 31.2 (8.19) | 30.5 (8.45) | 30.0 (6.93) | 28.5 (7.26) |
|  | [18.5, 25) | 9718 (24.9) | 9366 (24.9) | 352 (24.9) | 57 (17.5) | 37 (21.5) | 8 (14.3) | 3826 (20.6) |
|  | [25, 30) | 571 (1.5) | 559 (1.5) | 12 (0.9) | 4 (1.2) | 4 (2.3) | 1 (1.8) | 243 (1.3) |
|  | >=30 | 10187 (26.1) | 9799 (26.1) | 388 (27.5) | 103 (31.6) | 56 (32.6) | 23 (41.1) | 3781 (20.4) |
| **Ever-Smoker** | | 14197 (36.4) | 13773 (36.7) | 424 (30.0) | 134 (41.1) | 73 (42.4) | 22 (39.3) | 5378 (29.0) |
| **Smoking Status** | Never | 22832 (58.6) | 21978 (58.5) | 854 (60.5) | 161 (49.4) | 78 (45.3) | 18 (32.1) | 10763 (58.1) |
|  | Past | 10999 (28.2) | 10630 (28.3) | 369 (26.2) | 120 (36.8) | 66 (38.4) | 21 (37.5) | 3707 (20.0) |
|  | Current | 3198 (8.2) | 3143 (8.4) | 55 (3.9) | 14 (4.3) | 7 (4.1) | 1 (1.8) | 1671 (9.0) |
| **Alcohol Consumption** | | 20623 (52.9) | 19894 (53.0) | 729 (51.7) | 151 (46.3) | 78 (45.3) | 22 (39.3) | 6718 (36.3) |
| **SES,**  **mean (SD)** | NDI | 0.088 (0.06) | 0.088 (0.06) | 0.088 (0.06) | 0.095 (0.058) | 0.097 (0.058) | 0.096 (0.050) | 0.095 (0.064) |
|  | Population density (people/mi^2^) | 2110 (2260) | 2090 (2260) | 2410 (2250) | 2710 (2230) | 2900 (2340) | 2880 (1950) | 2060 (2320) |
| **Comorbidity Score,**  **mean (SD)** | | 2.29 (1.53) | 2.29 (1.53) | 2.26 (1.48) | 3.10 (1.64) | 3.34 (1.67) | 3.93 (1.62) | 1.39 (1.25) |

| **Black patients** | | **Total Tested for COVID-19** | | | | | | **Matched Comparison Group**  **(n=2165)** |
| --- | --- | --- | --- | --- | --- | --- | --- | --- |
|  |  | **Overall**  **(n=5763)** | **Negative**  **(n=5117)** | **Tested Positive** | | | |  |
|  |  |  |  | **Overall**  **(n=646)** | **Hospitalized**  **(n=265)** | **ICU**  **(n=139)** | **Deceased**  **(n=42)** |  |
| **Variable** | | n (%) |  |  |  |  |  |  |
| **Age**  **(in years)** | mean (SD) | 42.8 (21.7) | 42.1 (22.0) | 48.6 (18.6) | 56.8 (17.1) | 57.8 (17.0) | 67.2 (11.4) | 36.8 (23.2) |
|  | median [IQR] | 44.0 [34.0] | 43.0 [35.0] | 49.0 [27.0] | 58.0 [24.0] | 59.0 [22.5] | 66.5 [17.5] | 34.0 [38.0] |
| **Age Range** | [0,18) | 697 (12.1) | 675 (13.2) | 22 (3.4) | 5 (1.9) | 4 (2.9) | 0 (0) | 547 (25.3) |
|  | [18,35) | 1487 (25.8) | 1355 (26.5) | 132 (20.4) | 21 (7.9) | 11 (7.9) | 0 (0) | 541 (25.0) |
|  | [35,50) | 1173 (20.4) | 1001 (19.6) | 172 (26.6) | 63 (23.8) | 23 (16.5) | 3 (7.1) | 376 (17.4) |
|  | [50,65) | 1340 (23.3) | 1157 (22.6) | 183 (28.3) | 83 (31.3) | 50 (36.0) | 15 (35.7) | 384 (17.7) |
|  | [65,80) | 890 (15.4) | 784 (15.3) | 106 (16.4) | 67 (25.3) | 39 (28.1) | 15 (35.7) | 249 (11.5) |
|  | [80,100) | 176 (3.1) | 145 (2.8) | 31 (4.8) | 26 (9.8) | 12 (8.6) | 9 (21.4) | 68 (3.1) |
| **Male Sex** | | 2336 (40.5) | 2067 (40.4) | 269 (41.6) | 140 (52.8) | 81 (58.3) | 25 (59.5) | 1011 (46.7) |
| **Primary Care in MM** | | 3284 (57.0) | 2973 (58.1) | 311 (48.1) | 83 (31.3) | 34 (24.5) | 9 (21.4) | 431 (19.9) |
| **BMI, mean (SD)** | | 31.5 (8.80) | 31.1 (8.58) | 34.0 (9.94) | 34.6 (12.1) | 35.4 (14.3) | 33.1 (6.55) | 30.7 (8.31) |
| **BMI Range** | <18.5 | 996 (17.3) | 940 (18.4) | 56 (8.7) | 20 (7.5) | 13 (9.4) | 3 (7.1) | 298 (13.8) |
|  | [18.5, 25) | 100 (1.7) | 92 (1.8) | 8 (1.2) | 5 (1.9) | 0 (0) | 0 (0) | 22 (1.0) |
|  | [25, 30) | 1327 (23.0) | 1183 (23.1) | 144 (22.3) | 70 (26.4) | 36 (25.9) | 13 (31.0) | 357 (16.5) |
|  | >=30 | 2421 (42.0) | 2048 (40.0) | 373 (57.7) | 160 (60.4) | 84 (60.4) | 26 (61.9) | 583 (26.9) |
| **Ever-Smoker** | | 2011 (34.9) | 1831 (35.8) | 180 (27.9) | 83 (31.3) | 43 (30.9) | 19 (45.2) | 517 (23.9) |
| **Smoking Status** | Never | 3367 (58.4) | 2988 (58.4) | 379 (58.7) | 143 (54.0) | 62 (44.6) | 8 (19.0) | 1312 (60.6) |
|  | Past | 1350 (23.4) | 1200 (23.5) | 150 (23.2) | 77 (29.1) | 41 (29.5) | 19 (45.2) | 304 (14.0) |
|  | Current | 661 (11.5) | 631 (12.3) | 30 (4.6) | 6 (2.3) | 2 (1.4) | 0 (0) | 213 (9.8) |
| **Alcohol Consumption** | | 2393 (41.5) | 2134 (41.7) | 259 (40.1) | 81 (30.6) | 37 (26.6) | 12 (28.6) | 575 (26.6) |
| **SES, mean (SD)** | NDI | 0.177 (0.104) | 0.176 (0.104) | 0.187 (0.107) | 0.200 (0.112) | 0.210 (0.110) | 0.212 (0.095) | 0.189 (0.110) |
|  | Population density (people/mi^2^) | 3590 (2410) | 3550 (2410) | 3940 (2370) | 4440 (2480) | 4560 (2470) | 5050 (2260) | 3540 (2600) |
| **Comorbidity Score,**  **mean (SD)** | | 2.53 (1.61) | 2.52 (1.61) | 2.67 (1.64) | 3.18 (1.65) | 3.44 (1.63) | 4.27 (1.36) | 1.44 (1.26) |

Abbreviations: MM, Michigan Medicine; ICU, intensive care unit; BMI, body mass index; MM, Michigan Medicine; SES, social economics status; NDI, 2010 Neighborhood Socioeconomic Disadvantage Index.

**eTable 5.** Missingness of the Variables in the Full Cohort, White, and Black Patients

| Full Cohort | **COVID-19 Tested** | | | | | | **Comparison Group** |
| --- | --- | --- | --- | --- | --- | --- | --- |
|  | **Overall** | **Negative** | **COVID-19 Positive** | | | |  |
|  |  |  | **Overall** | **Hospitalized** | **ICU** | **Deceased** |  |
|  | **(n = 53853)** | **(n = 51271)** | **(n = 2582)** | **(n = 719)** | **(n = 377)** | **(n = 129)** | **(n = 29383)** |
| **Variable** | **n (%)** |  |  |  |  |  |  |
| **Age/Age Range** | 0 (0.0) | 0 (0.0) | 0 (0.0) | 0 (0.0) | 0 (0.0) | 0 (0.0) | 11 (0.0) |
| **Male Sex** | 2 (0.0) | 2 (0.0) | 0 (0.0) | 0 (0.0) | 0 (0.0) | 0 (0.0) | 19 (0.1) |
| **Primary Care in MM** | 0 (0.0) | 0 (0.0) | 0 (0.0) | 0 (0.0) | 0 (0.0) | 0 (0.0) | 0 (0.0) |
| **BMI/BMI Range** | 10506 (19.5) | 10104 (19.7) | 402 (15.6) | 36 (5.0) | 16 (4.2) | 5 (3.9) | 13272 (45.2) |
| **Ever-Smoker/ Smoking Status** | 4773 (8.9) | 4362 (8.5) | 411 (15.9) | 108 (15.0) | 86 (22.8) | 52 (40.3) | 7598 (25.9) |
| **Alcohol Consumption** | 16022 (29.8) | 15141 (29.5) | 881 (34.1) | 306 (42.6) | 176 (46.7) | 72 (55.8) | 13866 (47.2) |
| **Race Ethnicity**^a^ | 0 (0.0) | 0 (0.0) | 0 (0.0) | 0 (0.0) | 0 (0.0) | 0 (0.0) | 0.0 (0.0) |
| **SES (NDI/Population density)** | 7712 (14.3) | 7430 (14.5) | 282 (10.9) | 57 (7.9) | 28 (7.4) | 15 (11.6) | 5329 (18.1) |
| **Comorbidity Score** | 5991 (11.1) | 5542 (10.8) | 449 (17.4) | 218 (30.3) | 126 (33.4) | 38 (29.5) | 1490 (5.1) |
| White | **(n = 38977)** | **(n = 37566)** | **(n = 1411)** | **(n = 326)** | **(n = 172)** | **(n = 56)** | **(n = 18529)** |
| **Age/Age Range** | 0 (0) | 0 (0) | 0 (0) | 0 (0) | 0 (0) | 0 (0) | 0 (0) |
| **Male Sex** | 2 (0.0) | 2 (0.0) | 0 (0) | 0 (0) | 0 (0) | 0 (0) | 2 (0.0) |
| **Primary Care in MM** | 0 (0) | 0 (0) | 0 (0) | 0 (0) | 0 (0) | 0 (0) | 0 (0) |
| **BMI/BMI Range** | 6415 (16.5) | 6243 (16.6) | 172 (12.2) | 14 (4.3) | 7 (4.1) | 2 (3.6) | 6588 (35.6) |
| **Ever-Smoker/ Smoking Status** | 1948 (5.0) | 1815 (4.8) | 133 (9.4) | 31 (9.5) | 21 (12.2) | 16 (28.6) | 2388 (12.9) |
| **Alcohol Consumption** | 9884 (25.4) | 9519 (25.3) | 365 (25.9) | 108 (33.1) | 55 (32.0) | 25 (44.6) | 6812 (36.8) |
| **SES (NDI/Population density)** | 4687 (12.0) | 4553 (12.1) | 134 (9.5) | 36 (11.0) | 18 (10.5) | 9 (16.1) | 2970 (16.0) |
| **Comorbidity Score** | 2958 (7.6) | 2819 (7.5) | 139 (9.9) | 70 (21.5) | 40 (23.3) | 16 (28.6) | 501 (2.7) |
| Black | **(n = 5763)** | **(n = 5117)** | **(n = 646)** | **(n = 265)** | **(n = 139)** | **(n = 42)** | **(n = 2165)** |
| **Age/Age Range** | 0 (0) | 0 (0) | 0 (0) | 0 (0) | 0 (0) | 0 (0) | 0 (0) |
| **Male Sex** | 0 (0) | 0 (0) | 0 (0) | 0 (0) | 0 (0) | 0 (0) | 0 (0) |
| **Primary Care in MM** | 0 (0) | 0 (0) | 0 (0) | 0 (0) | 0 (0) | 0 (0) | 0 (0) |
| **BMI/BMI Range** | 919 (15.9) | 854 (16.7) | 65 (10.1) | 10 (3.8) | 6 (4.3) | 0 (0) | 905 (41.8) |
| **Ever-Smoker/ Smoking Status** | 385 (6.7) | 298 (5.8) | 87 (13.5) | 39 (14.7) | 34 (24.5) | 15 (35.7) | 336 (15.5) |
| **Alcohol Consumption** | 1665 (28.9) | 1435 (28.0) | 230 (35.6) | 123 (46.4) | 75 (54.0) | 22 (52.4) | 878 (40.6) |
| **SES (NDI/Population density)** | 720 (12.5) | 674 (13.2) | 46 (7.1) | 10 (3.8) | 4 (2.9) | 1 (2.4) | 274 (12.7) |
| **Comorbidity Score** | 656 (11.4) | 517 (10.1) | 139 (21.5) | 90 (34.0) | 53 (38.1) | 12 (28.6) | 75 (3.5) |

Abbreviations: MM, Michigan Medicine; ICU, intensive care unit; BMI, body mass index; MM, Michigan Medicine; SES, social economics status; NDI, 2010 Neighborhood Socioeconomic Disadvantage Index.

^a^ In the main analyses unknown race and/or ethnicity were combined into the category "Unknown race/ethnicity" to retain sample size

**eTable 6. Odds Ratios of COVID-19 Outcomes from Logistic Regression, Stratified by Quarters**

| **Tested (1) vs. Comparison Group (0)** | | | **Full Cohort**  (n_0_=22988, n_1_=43821) | **Quarter 1**  (n_0_=22988, n_1_=1896) | **Quarter 2**  (n_0_=22988, n_1_=19644) | **Quarter 3**  (n_0_=22988, n_1_=21759) |
| --- | --- | --- | --- | --- | --- | --- |
| **Variable** | | | **OR (95% CI)** |  |  |  |
| **Age (unit: 10-year)** | | | 0.93 (0.92, 0.94) | 0.88 (0.86, 0.90) | 0.96 (0.96, 0.97) | 0.90 (0.89, 0.91) |
| **Age Range**  REF: [18,35] | | [0,18] | 1.97 (1.81, 2.15) | 3.52 (2.68, 4.62) | 1.62 (1.46, 1.81) | 2.05 (1.86, 2.26) |
|  |  | [35,50] | 2.66 (2.32, 3.06) | 5.92 (3.90, 8.99) | 2.24 (1.89, 2.66) | 2.69 (2.29, 3.16) |
|  |  | [50,65] | 2.94 (2.43, 3.57) | 4.05 (2.29, 7.17) | 2.43 (1.91, 3.07) | 3.16 (2.53, 3.95) |
|  |  | [65,80] | 3.15 (2.47, 4.01) | 2.48 (1.20, 5.11) | 2.58 (1.92, 3.48) | 3.48 (2.63, 4.61) |
|  |  | [80,100] | 2.76 (2.04, 3.72) | 2.96 (1.21, 7.24) | 2.38 (1.65, 3.43) | 2.79 (1.97, 3.95) |
| **Male Sex** | | | 0.86 (0.83, 0.89) | 0.64 (0.58, 0.71) | 0.86 (0.82, 0.90) | 0.88 (0.85, 0.92) |
| **BMI** | | | 1.00 (1.00, 1.01) | 1.01 (1.00, 1.01) | 1.01 (1.00, 1.01) | 0.999 (0.996, 1.00) |
| **BMI Range**  REF: [18.5,25] | | <18.5 | 0.94 (0.81, 1.08) | 0.87 (0.58, 1.31) | 1.02 (0.86, 1.21) | 0.92 (0.77, 1.09) |
|  |  | [25,30] | 1.07 (1.02, 1.13) | 1.10 (0.96, 1.27) | 1.11 (1.05, 1.18) | 1.05 (0.99, 1.11) |
|  |  | >=30 | 1.07 (1.01, 1.12) | 1.23 (1.08, 1.41) | 1.12 (1.05, 1.19) | 1.00 (0.95, 1.06) |
| **Ever-Smoker** | | |  | 1.13 (1.08, 1.17) | 1.13 (1.01, 1.27) | 1.18 (1.13, 1.24) |
| **Smoking Status**  REF: Never-Smoker | | Past-Smoker | 1.21 (1.15, 1.27) | 1.25 (1.10, 1.42) | 1.26 (1.19, 1.33) | 1.17 (1.11, 1.24) |
|  |  | Current-Smoker | 0.95 (0.89, 1.02) | 0.87 (0.71, 1.06) | 1.02 (0.95, 1.11) | 0.93 (0.86, 1.00) |
| **Alcohol Consumption** | | |  | 1.96 (1.87, 2.05) | 2.16 (1.91, 2.44) | 1.89 (1.80, 2.00) |
| **Race/Ethnicity**  REF: White | | Black | 1.22 (1.14, 1.30) | 2.15 (1.85, 2.5) | 1.30 (1.21, 1.40) | 1.05 (0.98, 1.14) |
|  |  | Other / Known Ethnicity | 0.92 (0.87, 0.98) | 1.16 (0.99, 1.37) | 0.90 (0.84, 0.97) | 0.91 (0.85, 0.99) |
|  |  | Other / Unknown Ethnicity | 0.39 (0.36, 0.41) | 0.33 (0.26, 0.43) | 0.34 (0.32, 0.37) | 0.44 (0.41, 0.47) |
| **SES** | | Population density (1000-people/mi^2^) | 1.03 (1.03, 1.04) | 1.06 (1.04, 1.08) | 1.02 (1.01, 1.03) | 1.04 (1.03, 1.05) |
|  |  | NDI | 0.10 (0.07, 0.13) | 0.05 (0.02, 0.10) | 0.25 (0.18, 0.34) | 0.04 (0.03, 0.06) |
| **Comorbidity Score** | | |  | 1.75 (1.72, 1.77) | 1.89 (1.82, 1.96) | 1.77 (1.74, 1.80) |
| **Comorbidities** | | Respiratory | 2.97 (2.87, 3.07) | 4.34 (3.86, 4.88) | 2.87 (2.76, 3.00) | 2.93 (2.82, 3.05) |
|  |  | Circulatory | 3.02 (2.91, 3.13) | 3.58 (3.20, 4.01) | 3.20 (3.05, 3.34) | 2.78 (2.66, 2.90) |
|  |  | Any Cancer | 1.73 (1.66, 1.81) | 1.72 (1.52, 1.94) | 1.97 (1.87, 2.07) | 1.60 (1.52, 1.69) |
|  |  | Type 2 Diabetes | 2.01 (1.90, 2.12) | 2.11 (1.83, 2.42) | 2.09 (1.97, 2.23) | 1.86 (1.74, 1.98) |
|  |  | Kidney | 3.27 (3.06, 3.50) | 3.81 (3.30, 4.41) | 3.85 (3.58, 4.14) | 2.69 (2.49, 2.90) |
|  |  | Liver | 3.58 (3.27, 3.91) | 4.23 (3.55, 5.04) | 3.90 (3.55, 4.30) | 3.23 (2.93, 3.56) |
|  |  | Autoimmune | 2.06 (1.95, 2.18) | 2.85 (2.51, 3.23) | 2.11 (1.98, 2.25) | 1.91 (1.79, 2.03) |
| **Positive (1) vs Comparison Group (0)** | | | **Full Cohort**  (n0=29383, n1=2582) | **Quarter 1**  (n0=29383, n1=4832) | **Quarter 2**  (n0=29383, n1=823) | **Quarter 3**  (n0=29383, n1=683) |
| **Variable** | | | **OR (95% CI)** |  |  |  |
| **Age (unit: 10-year)** | | | 0.97 (0.944, 0.99) | 1.05 (0.998, 1.11) | 1.03 (0.99, 1.08) | 0.853 (0.817, 0.89) |
| **Age Range**  REF: [18,35) | [0,18) | | 0.31 (0.244, 0.38) | 0.0422 (0.012, 0.149) | 0.371 (0.239, 0.577) | 0.399 (0.294, 0.54) |
|  | [35,50) | | 1.14 (0.986, 1.32) | 1.5 (1.09, 2.06) | 1.36 (1.02, 1.8) | 0.654 (0.509, 0.84) |
|  | [50,65) | | 0.848 (0.734, 0.981) | 1.21 (0.877, 1.66) | 1.18 (0.894, 1.55) | 0.431 (0.333, 0.559) |
|  | [65,80) | | 0.525 (0.444, 0.62) | 0.6 (0.406, 0.886) | 1.02 (0.762, 1.36) | 0.251 (0.183, 0.343) |
|  | [80,100) | | 0.392 (0.305, 0.503) | 0.711 (0.425, 1.19) | 0.758 (0.505, 1.14) | 0.144 (0.0814, 0.255) |
| **Male Sex** | | | 0.871 (0.791, 0.96) | 0.895 (0.72, 1.11) | 0.898 (0.756, 1.07) | 0.826 (0.692, 0.985) |
| **BMI** | | | 1.02 (1.01, 1.02) | 1.02 (1.01, 1.03) | 1.02 (1.01, 1.03) | 1.01 (0.998, 1.02) |
| **BMI Range**  REF: [18.5,25) | <18.5 | | 0.705 (0.437, 1.14) | 0.577 (0.162, 2.06) | 0.571 (0.219, 1.49) | 1.12 (0.57, 2.2) |
|  | [25,30) | | 1.31 (1.14, 1.52) | 1.59 (1.15, 2.2) | 1.16 (0.897, 1.5) | 1.3 (1.01, 1.68) |
|  | >=30 | | 1.5 (1.31, 1.72) | 1.94 (1.43, 2.63) | 1.45 (1.14, 1.83) | 1.39 (1.08, 1.78) |
| **Ever-Smoker** | | | 0.803 (0.714, 0.902) | 0.777 (0.604, 0.999) | 0.914 (0.749, 1.12) | 0.691 (0.549, 0.871) |
| **Smoking Status**  REF: Never-Smoker | Past-Smoker | | 0.954 (0.841, 1.08) | 0.979 (0.751, 1.28) | 1.02 (0.826, 1.27) | 0.799 (0.621, 1.03) |
|  | Current-Smoker | | 0.447 (0.353, 0.565) | 0.29 (0.155, 0.543) | 0.647 (0.451, 0.929) | 0.458 (0.294, 0.713) |
| **Alcohol Consumption** | | | 1.69 (1.5, 1.9) | 2.14 (1.66, 2.77) | 1.31 (1.07, 1.61) | 1.5 (1.21, 1.86) |
| **Race/Ethnicity**  REF: White | Black | | 3.21 (2.8, 3.67) | 7.94 (6.1, 10.3) | 3.5 (2.79, 4.39) | 1.59 (1.2, 2.12) |
|  | Other / Known Ethnicity | | 1.25 (1.06, 1.48) | 2.02 (1.41, 2.91) | 1.4 (1.03, 1.89) | 1.03 (0.771, 1.39) |
|  | Other / Unknown Ethnicity | | 0.611 (0.504, 0.742) | 0.426 (0.239, 0.762) | 0.66 (0.464, 0.937) | 0.569 (0.4, 0.808) |
| **SES** | Population density (1000-people/mi^2^) | | 0.192 (0.0975, 0.379) | 0.0809 (0.0193, 0.339) | 1.07 (0.362, 3.16) | 0.062 (0.0155, 0.247) |
|  | NDI | | 1.07 (1.05, 1.1) | 1.08 (1.04, 1.12) | 1.09 (1.06, 1.12) | 1.03 (0.997, 1.07) |
| **Comorbidity Score** | | | 1.71 (1.64, 1.77) | 1.59 (1.47, 1.72) | 1.8 (1.69, 1.91) | 1.6 (1.49, 1.71) |
| **Comorbidities** | Respiratory | | 3.74 (3.34, 4.17) | 3.36 (2.61, 4.32) | 3.27 (2.69, 3.97) | 3.27 (2.67, 3.99) |
|  | Circulatory | | 3.09 (2.77, 3.45) | 2.46 (1.91, 3.17) | 3.56 (2.89, 4.4) | 2.46 (2.03, 2.99) |
|  | Any Cancer | | 1.19 (1.05, 1.34) | 1.18 (0.899, 1.55) | 1.58 (1.29, 1.94) | 1.19 (0.929, 1.52) |
|  | Type 2 Diabetes | | 2.26 (1.98, 2.57) | 1.73 (1.3, 2.31) | 2.86 (2.32, 3.52) | 1.71 (1.28, 2.27) |
|  | Kidney | | 2.96 (2.56, 3.43) | 2.39 (1.75, 3.28) | 4.05 (3.25, 5.05) | 2.28 (1.67, 3.12) |
|  | Liver | | 3.04 (2.52, 3.67) | 3.11 (2.1, 4.62) | 3.58 (2.67, 4.8) | 3.2 (2.27, 4.53) |
|  | Autoimmune | | 2.33 (2.05, 2.65) | 2.72 (2.08, 3.55) | 2.39 (1.92, 2.97) | 1.83 (1.4, 2.38) |
| **Hospitalization (1) vs Not (0)** | | | **Full Cohort**  (n0=1852, n1=719) | **Quarter 1**  (n0=282, n1=200) | **Quarter 2**  (n0=454, n1=363) | **Quarter 3**  (n0=597, n1=86) |
| **Age (unit: 10-year)** | | | 1.46 (1.36, 1.56) | 2.09 (1.7, 2.57) | 1.20 (1.09, 1.33) | 1.55 (1.32, 1.83) |
| **Age Range**  REF: [18,35) | [0,18) | | 1.24 (0.604, 2.56) | NA (NA, NA) | 1.99 (0.75, 5.25) | 0.195 (0.012, 3.24) |
|  | [35,50) | | 1.51 (1.00, 2.28) | 2.41 (0.952, 6.12) | 1.17 (0.61, 2.22) | 2.09 (0.85, 5.14) |
|  | [50,65) | | 2.62 (1.78, 3.85) | 5.7 (2.28, 14.3) | 1.58 (0.87, 2.87) | 2.40 (0.97, 5.93) |
|  | [65,80) | | 4.62 (3.07, 6.95) | 11 (3.88, 31.1) | 2.18 (1.18, 4.04) | 7.58 (3.09, 18.6) |
|  | [80,100) | | 13.3 (7.51, 23.4) | 113 (16.6, 774) | 5.80 (2.37, 14.2) | 10.4 (2.45, 44.2) |
| **Male Sex** | | | 1.84 (1.46, 2.31) | 2.36 (1.37, 4.08) | 1.88 (1.31, 2.70) | 1.58 (0.905, 2.75) |
| **BMI** | | | 1.02 (1.00, 1.04) | 1.02 (0.983, 1.06) | 1.01 (0.987, 1.04) | 1.04 (1, 1.08) |
| **BMI Range**  REF: [18.5,25) | <18.5 | | 3.61 (1.17, 11.2) | 6.64 (0.153, 289) | 1.27 (0.127, 12.7) | 14.9 (2.61, 85.5) |
|  | [25,30) | | 1.47 (1.02, 2.14) | 1.95 (0.767, 4.97) | 1.11 (0.634, 1.93) | 2.33 (0.839, 6.45) |
|  | >=30 | | 1.60 (1.12, 2.27) | 2.23 (0.879, 5.66) | 1.16 (0.696, 1.92) | 3.41 (1.3, 8.94) |
| **Ever-Smoker** | | | 1.09 (0.85, 1.40) | 1.32 (0.735, 2.37) | 0.76 (0.50, 1.14) | 1.61 (0.887, 2.92) |
| **Smoking Status**  REF: Never-Smoker | Past-Smoker | | 1.15 (0.88, 1.49) | 1.46 (0.797, 2.67) | 0.80 (0.52, 1.23) | 1.70 (0.907, 3.18) |
|  | Current-Smoker | | 0.78 (0.43, 1.43) | 0.547 (0.0978, 3.06) | 0.62 (0.277, 1.4) | 1.31 (0.357, 4.84) |
| **Alcohol Consumption** | | | 0.77 (0.59, 1.01) | 0.943 (0.498, 1.79) | 0.66 (0.43, 1.02) | 0.696 (0.38, 1.29) |
| **Race/Ethnicity**  REF: White | Black | | 1.85 (1.39, 2.47) | 1.43 (0.752, 2.71) | 1.25 (0.79, 1.98) | 1.76 (0.81, 3.85) |
|  | Other / Known Ethnicity | | 1.15 (0.75, 1.75) | 1.31 (0.513, 3.33) | 0.94 (0.49, 1.82) | 0.97 (0.35, 2.72) |
|  | Other / Unknown Ethnicity | | 1.11 (0.69, 1.78) | 0.922 (0.205, 4.15) | 0.83 (0.38, 1.83) | 0.07 (0.0036, 1.37) |
| **SES** | Population density (1000-people/mi^2^) | | 11.7 (3.08, 44.4) | 664 (20.4, 21600) | 1.72 (0.22, 13.5) | 3.69 (0.103, 132) |
|  | NDI | | 1.07 (1.02, 1.13) | 1.05 (0.93, 1.2) | 1.09 (1.00, 1.18) | 0.97 (0.85, 1.12) |
| **Comorbidity Score** | | | 1.24 (1.15, 1.35) | 1.13 (0.94, 1.36) | 1.28 (1.14, 1.45) | 1.29 (1.06, 1.57) |
| **Comorbidities** | Respiratory | | 1.14 (0.86, 1.52) | 0.91 (0.47, 1.74) | 0.991 (0.656, 1.5) | 1.88 (0.898, 3.92) |
|  | Circulatory | | 1.58 (1.16, 2.15) | 1.08 (0.57, 2.03) | 2.01 (1.24, 3.27) | 1.67 (0.79, 3.54) |
|  | Any Cancer | | 1.17 (0.90, 1.53) | 1.19 (0.62, 2.26) | 1.32 (0.891, 1.96) | 0.93 (0.481, 1.8) |
|  | Type 2 Diabetes | | 1.79 (1.39, 2.32) | 1.39 (0.71, 2.74) | 1.91 (1.27, 2.88) | 1.79 (0.911, 3.5) |
|  | Kidney | | 3.20 (2.41, 4.24) | 3.04 (1.40, 6.60) | 3.43 (2.21, 5.31) | 2.99 (1.46, 6.12) |
|  | Liver | | 1.24 (0.86, 1.80) | 0.44 (0.16, 1.23) | 1.41 (0.792, 2.5) | 1.77 (0.802, 3.9) |
|  | Autoimmune | | 1.06 (0.80, 1.41) | 1.90 (0.996, 3.62) | 1.21 (0.779, 1.88) | 1.22 (0.607, 2.47) |
| **ICU (1) vs Not (0)** | | | **Full Cohort**  (n0=2194, n1=377) | **Quarter 1**  (n0=364, n1=118) | **Quarter 2**  (n0=619, n1=198) | **Quarter 3**  (n0=647, n1=36) |
| **Age (unit: 10-year)** | | | 1.34 (1.23, 1.47) | 2.14 (1.7, 2.71) | 1.07 (0.95, 1.21) | 1.27 (1.01, 1.59) |
| **Age Range**  REF: [18,35) | [0,18) | | 2.21 (0.906, 5.38) | NA (NA, NA) | 2.34 (0.77, 7.14) | 0.52 (0.0297, 9.19) |
|  | [35,50) | | 1.42 (0.778, 2.58) | 4 (0.686, 23.4) | 0.76 (0.34, 1.73) | 2.81 (0.76, 10.4) |
|  | [50,65) | | 2.56 (1.48, 4.44) | 16.4 (3.03, 88.5) | 1.15 (0.55, 2.37) | 1.02 (0.24, 4.46) |
|  | [65,80) | | 4.39 (2.51, 7.7) | 35.8 (6.2, 207) | 1.66 (0.81, 3.44) | 4.67 (1.22, 17.8) |
|  | [80,100) | | 5.39 (2.67, 10.9) | 69.2 (10.5, 457) | 1.27 (0.45, 3.55) | 2.24 (0.236, 21.2) |
| **Male Sex** | | | 2.33 (1.74, 3.13) | 2.41 (1.29, 4.51) | 1.97 (1.28, 3.01) | 3.82 (1.62, 9.03) |
| **BMI** | | | 1.01 (0.99, 1.03) | 1.03 (0.986, 1.08) | 0.998 (0.97, 1.03) | 1.01 (0.953, 1.07) |
| **BMI Range**  REF: [18.5,25) | <18.5 | | 2.96 (0.76, 11.6) | 13.6 (0.201, 928) | NA (NA, NA) | 12.9 (1.92, 87.1) |
|  | [25,30) | | 1.17 (0.73, 1.86) | 1.71 (0.574, 5.11) | 1.03 (0.54, 1.97) | 1.03 (0.28, 3.8) |
|  | >=30 | | 1.08 (0.69, 1.68) | 1.92 (0.642, 5.75) | 0.83 (0.46, 1.50) | 1.01 (0.28, 3.58) |
| **Ever-Smoker** | | | 1.29 (0.94, 1.77) | 2.22 (1.17, 4.21) | 0.84 (0.52, 1.36) | 1.61 (0.69, 3.74) |
| **Smoking Status**  REF: Never-Smoker | Past-Smoker | | 1.35 (0.97, 1.87) | 2.39 (1.24, 4.6) | 0.86 (0.52, 1.43) | 1.84 (0.76, 4.43) |
|  | Current-Smoker | | 0.997 (0.46, 2.16) | 1.11 (0.152, 8.09) | 0.798 (0.31, 2.07) | 0.895 (0.13, 6.3) |
| **Alcohol Consumption** | | | 0.81 (0.57, 1.15) | 1.20 (0.57, 2.49) | 0.74 (0.449, 1.22) | 0.697 (0.28, 1.75) |
| **Race/Ethnicity**  REF: White | Black | | 1.33 (0.93, 1.90) | 0.99 (0.47, 2.06) | 0.94 (0.55, 1.6) | 0.73 (0.22, 2.45) |
|  | Other / Known Ethnicity | | 0.82 (0.45, 1.48) | 0.998 (0.31, 3.23) | 0.59 (0.24, 1.45) | 0.63 (0.15, 2.66) |
|  | Other / Unknown Ethnicity | | 1.43 (0.81, 2.52) | 1.12 (0.198, 6.27) | 1.40 (0.59, 3.36) | 0.24 (0.013, 4.46) |
| **SES** | Population density (1000-people/mi^2^) | | 20.7 (4.17, 102) | 236 (4.99, 11200) | 3.07 (0.293, 32.2) | 76.8 (0.67, 8770) |
|  | NDI | | 1.10 (1.03, 1.17) | 1.06 (0.919, 1.22) | 1.11 (1.02, 1.21) | 1.08 (0.91, 1.27) |
| **Comorbidity Score** | | | 1.34 (1.21, 1.48) | 1.04 (0.85, 1.27) | 1.4 (1.21, 1.62) | 1.56 (1.18, 2.04) |
| **Comorbidities** | Respiratory | | 1.36 (0.94, 1.97) | 0.65 (0.31, 1.36) | 1.66 (0.99, 2.79) | 2.46 (0.83, 7.26) |
|  | Circulatory | | 1.62 (1.07, 2.45) | 0.97 (0.46, 2.08) | 2.62 (1.37, 5.00) | 1.24 (0.44, 3.49) |
|  | Any Cancer | | 1.32 (0.96, 1.83) | 0.995 (0.49, 2.00) | 1.45 (0.93, 2.29) | 1.13 (0.45, 2.86) |
|  | Type 2 Diabetes | | 1.89 (1.39, 2.59) | 0.79 (0.38, 1.65) | 2.01 (1.27, 3.2) | 3.30 (1.39, 7.81) |
|  | Kidney | | 3.64 (2.64, 5.02) | 2.20 (1.05, 4.60) | 3.89 (2.42, 6.25) | 2.81 (1.14, 6.93) |
|  | Liver | | 1.34 (0.86, 2.09) | 0.66 (0.22, 2.00) | 0.98 (0.499, 1.91) | 5.81 (2.31, 14.6) |
|  | Autoimmune | | 1.30 (0.93, 1.84) | 1.88 (0.93, 3.82) | 1.43 (0.87, 2.35) | 1.68 (0.64, 4.40) |
| **Deceased (1) vs Alive (0)** | | | **Full Cohort**  (n0=2453, n1=129) | **Quarter 1**  (n0=448, n1=35) | **Quarter 2**  (n0=753, n1=70) | **Quarter 3**  (n0=679, n1=4) |
| **Age (unit: 10-year)** | | | 2.1 (1.74, 2.52) | 2.42 (1.62, 3.63) | 1.78 (1.39, 2.27) | 1.5 (0.84, 2.68) |
| **Age Range**  REF: [18,35) | [0,18) | | 1.09 (0.0532, 22.1) | NA (NA, NA) | 0.89 (0.04, 19.7) | 0.266 (0.0065, 10.9) |
|  | [35,50) | | 1.74 (0.411, 7.38) | NA (NA, NA) | 0.86 (0.16, 4.60) | 0.136 (0.000334, 55.1) |
|  | [50,65) | | 2.95 (0.772, 11.3) | NA (NA, NA) | 1.50 (0.36, 6.29) | 0.0509 (1.34e-05, 193) |
|  | [65,80) | | 7.14 (1.91, 26.7) | NA (NA, NA) | 2.85 (0.72, 11.2) | 0.263 (9.11e-06, 7590) |
|  | [80,100) | | 36.5 (9.49, 140) | NA (NA, NA) | 15.1 (3.45, 65.8) | 0.038 (1.68e-07, 8560) |
| **Male Sex** | | | 1.93 (1.19, 3.13) | 2.64 (0.88, 7.93) | 2.30 (1.18, 4.52) | 3.25 (0.38, 27.8) |
| **BMI** | | | 1.02 (0.985, 1.05) | 0.982 (0.899, 1.07) | 1.03 (0.98, 1.07) | 1.04 (0.90, 1.19) |
| **BMI Range**  REF: [18.5,25) | <18.5 | | 6.34 (0.652, 61.6) | 231 (2.89, 18500) | NA (NA, NA) | 14.7 (0.39, 550) |
|  | [25,30) | | 1.54 (0.655, 3.62) | 0.687 (0.0887, 5.33) | 0.769 (0.257, 2.3) | 0.896 (0.068, 11.7) |
|  | >=30 | | 2.08 (0.906, 4.79) | 0.848 (0.109, 6.57) | 1.93 (0.745, 5.02) | 0.73 (0.055, 9.64) |
| **Ever-Smoker** | | | 1.45 (0.82, 2.58) | 0.994 (0.328, 3.01) | 1.23 (0.569, 2.66) | 1.77 (0.29, 10.9) |
| **Smoking Status**  REF: Never-Smoker | Past-Smoker | | 1.50 (0.843, 2.68) | 1.04 (0.338, 3.17) | 1.29 (0.588, 2.82) | 2.10 (0.31, 14.1) |
|  | Current-Smoker | | 1.09 (0.182, 6.46) | 2.32 (0.0947, 56.9) | 1.02 (0.162, 6.45) | 3.82 (0.22, 67.5) |
| **Alcohol Consumption** | | | 1.05 (0.562, 1.95) | 1.79 (0.523, 6.15) | 1.03 (0.457, 2.31) | 0.16 (0.017, 1.45) |
| **Race/Ethnicity**  REF: White | Black | | 1.46 (0.813, 2.64) | 1.72 (0.508, 5.81) | 0.879 (0.374, 2.07) | 1.66 (0.107, 25.7) |
|  | Other / Known Ethnicity | | 1.26 (0.479, 3.32) | 0.834 (0.0378, 18.4) | 1.61 (0.529, 4.89) | 3.46 (0.3, 40) |
|  | Other / Unknown Ethnicity | | 5.44 (2.63, 11.3) | 1.85 (0.0659, 51.9) | 3.74 (1.26, 11.1) | 2.93 (0.14, 62.5) |
| **SES** | Population density (1000-people/mi^2^) | | 110 (9.92, 1230) | 333 (0.92, 121000) | 14.1 (0.47, 428) | 20.2 (0.0005, 798000) |
|  | NDI | | 1.11 (1.01, 1.23) | 1.24 (0.97, 1.58) | 1.10 (0.96, 1.26) | 0.95 (0.64, 1.41) |
| **Comorbidity Score** | | | 1.52 (1.29, 1.79) | 1.37 (0.99, 1.89) | 1.43 (1.14, 1.78) | 1.49 (0.78, 2.83) |
| **Comorbidities** | Respiratory | | 2.50 (1.22, 5.13) | 0.62 (0.18, 2.16) | 3.35 (1.27, 8.85) | 1.51 (0.15, 15.3) |
|  | Circulatory | | 2.91 (1.09, 7.78) | 1.09 (0.26, 4.63) | 3.29 (0.87, 12.5) | 0.63 (0.06, 6.43) |
|  | Any Cancer | | 1.62 (0.99, 2.65) | 1.98 (0.72, 5.43) | 1.73 (0.87, 3.45) | 5.80 (0.57, 59.2) |
|  | Type 2 Diabetes | | 3.39 (2.07, 5.57) | 1.87 (0.64, 5.44) | 2.92 (1.47, 5.82) | 5.81 (0.75, 44.9) |
|  | Kidney | | 3.82 (2.31, 6.31) | 5.45 (1.82, 16.3) | 2.42 (1.22, 4.81) | 0.12 (0.005, 2.87) |
|  | Liver | | 1.67 (0.853, 3.26) | 1.15 (0.24, 5.53) | 1.10 (0.41, 2.97) | 15.3 (2.14, 110) |
|  | Autoimmune | | 0.93 (0.524, 1.67) | 1.54 (0.517, 4.6) | 0.85 (0.37, 1.94) | 0.82 (0.06, 10.5) |

Abbreviations: OR, odds ratio; ICU, intensive care unit; BMI, body mass index; NA, not applicable; REF, reference group; SES, social economics status; NDI, 2010 Neighborhood Socioeconomic Disadvantage Index; Quarter 1, March 10--March 31, 2020; Quarter 2, April 1—June 30, 2020; Quarter 3, July 1—September 2, 2020.

The model used was: $logit P\left( Y_{\mathrm{COVID}}=1|X, adjustment \right)=\beta_{0}+\beta_{X}X+\beta_{\mathrm{adjust}}\mathrm{adjustment}_{3}$. Here $Y_{\mathrm{COVID}}$ is various COVID-19 related outcomes under consideration (i.e., COVID-19 positive, hospitalization and ICU admission); $X$ is the variable/risk factor of interest; and $\mathrm{adjustment}_{3}$ is listed in eTable 1.

Note: green represents significant OR<1 (protective effect), orange represents significant OR>=1 (risk effect) under P<0.05.

**eTable 7. Proportion of Transferred Patients by Outcome and Testing Quarter**

| **Outcome** | **Transferred** | **Tested Positive** | | |  | **Diagnosed** | **Total** |
| --- | --- | --- | --- | --- | --- | --- | --- |
|  |  | **Q1** | **Q2** | **Q3** |  | **Unknown Quarter** |  |
| Tested Positive / Diagnosed for COVID | Yes | 30 | 93 | 11 |  | 35 | 169 |
|  | No | 453 | 730 | 672 |  | 558 | 2413 |
|  | % | 6.2% | 11.3% | 1.6% |  | 5.9% | 6.5% |
| Hospitalized | Yes | 30 | 93 | 11 |  | 35 | 169 |
|  | No | 170 | 270 | 75 |  | 35 | 550 |
|  | % | 15.0% | 25.6% | 12.8% |  | 50.0% | 23.5% |
| ICU | Yes | 27 | 84 | 9 |  | 21 | 141 |
|  | No | 91 | 114 | 27 |  | 4 | 236 |
|  | % | 22.9% | 42.4% | 25.0% |  | 84.0% | 37.4% |
| Deceased | Yes | 11 | 24 | 2 |  | 15 | 52 |
|  | No | 24 | 46 | 2 |  | 5 | 77 |
|  | % | 31.4% | 34.3% | 50.0% |  | 75.0% | 40.3% |

Abbreviations: ICU, intensive care unit; Q1, March 10 - March 31, 2020; Q2, April 1 - June 30, 2020; Q3, July 1 - September 2, 2020.

**eTable 8. Sensitivity Analysis Using Patients with Primary Care at Michigan Medicine, Stratified by Quarters.**

| **Hospitalization (1) vs Not (0)** | | **Full Cohort**  (n0=1025, n1=223) | **Quarter 1**  (n0=66, n1=175) | **Quarter 2**  (n0=254, n1=114) | **Quarter 3**  (n0=326, n1=39) |
| --- | --- | --- | --- | --- | --- |
| **Age (unit: 10-year)** | | 1.42 (1.28, 1.57) | 2.12 (1.58, 2.84) | 1.19 (1.03, 1.36) | 1.63 (1.30, 2.04) |
| **Age Range**  REF: [18,35) | [0,18) | 1.33 (0.50, 3.55) | NA (NA, NA) | 1.73 (0.498, 5.98) | 0.40 (0.02, 7.12) |
|  | [35,50) | 1.01 (0.58, 1.77) | 2.43 (0.668, 8.8) | 0.726 (0.327, 1.61) | 1.03 (0.248, 4.28) |
|  | [50,65) | 1.55 (0.93, 2.59) | 4.25 (1.21, 14.9) | 0.867 (0.413, 1.82) | 2.69 (0.83, 8.69) |
|  | [65,80) | 3.42 (1.97, 5.94) | 10.4 (2.43, 44.9) | 1.50 (0.677, 3.31) | 8.22 (2.53, 26.8) |
|  | [80,100) | 9.64 (4.53, 20.5) | 65.9 (7.34, 592) | 5.76 (1.72, 19.3) | 7.89 (1.22, 51) |
| **Male Sex** | | 1.66 (1.21, 2.29) | 2.24 (1.06, 4.74) | 1.59 (0.991, 2.56) | 1.68 (0.808, 3.5) |
| **BMI** | | 1.02 (1.00, 1.05) | 1.01 (0.958, 1.06) | 1.03 (0.993, 1.06) | 1.03 (0.979, 1.09) |
| **BMI Range**  REF: [18.5,25) | <18.5 | 2.25 (0.32, 15.9) | NA (NA, NA) | NA (NA, NA) | 5.04 (0.457, 55.6) |
|  | [25,30) | 1.41 (0.84, 2.35) | 1.85 (0.53, 6.43) | 1.46 (0.705, 3.04) | 1.57 (0.485, 5.12) |
|  | >=30 | 1.52 (0.93, 2.47) | 1.92 (0.553, 6.68) | 1.31 (0.663, 2.58) | 1.82 (0.586, 5.65) |
| **Ever-Smoker** | | 1.19 (0.85, 1.66) | 1.58 (0.731, 3.42) | 0.88 (0.525, 1.47) | 1.59 (0.747, 3.39) |
| **Smoking Status**  REF: Never-Smoker | Past-Smoker | 1.26 (0.89, 1.78) | 1.75 (0.791, 3.86) | 0.941 (0.547, 1.62) | 1.55 (0.702, 3.43) |
|  | Current-Smoker | 0.82 (0.37, 1.83) | 0.712 (0.0779, 6.51) | 0.647 (0.216, 1.94) | 2.21 (0.466, 10.5) |
| **Alcohol Consumption** | | 0.57 (0.40, 0.81) | 0.574 (0.26, 1.27) | 0.566 (0.332, 0.966) | 0.555 (0.253, 1.22) |
| **Race/Ethnicity**  REF: White | Black | 1.96 (1.32, 2.91) | 2.71 (1.13, 6.51) | 1.2 (0.665, 2.18) | 1.21 (0.397, 3.68) |
|  | Other / Known Ethnicity | 1.48 (0.87, 2.50) | 2.86 (0.886, 9.21) | 1.46 (0.674, 3.18) | 1.04 (0.278, 3.93) |
|  | Other / Unknown Ethnicity | 0.47 (0.15, 1.49) | 0.597 (0.0251, 14.2) | 0.976 (0.25, 3.81) | 0.132 (0.00555, 3.16) |
| **SES** | Population density (1000-people/mi^2^) | 1.02 (0.94, 1.10) | 0.979 (0.821, 1.17) | 1.03 (0.919, 1.15) | 0.918 (0.749, 1.12) |
|  | NDI | 4.47 (0.65, 30.9) | 196 (2.34, 16500) | 0.757 (0.0433, 13.2) | 3.77 (0.0325, 438) |
| **Comorbidity Score** | | 1.37 (1.22, 1.54) | 1.33 (1.02, 1.73) | 1.36 (1.15, 1.62) | 1.45 (1.12, 1.89) |
| **Comorbidities** | Respiratory | 0.94 (0.61, 1.43) | 2.55 (0.802, 8.11) | 0.709 (0.399, 1.26) | 2.18 (0.718, 6.62) |
|  | Circulatory | 1.71 (1.05, 2.79) | 1.44 (0.554, 3.77) | 2.41 (1.19, 4.89) | 1.39 (0.465, 4.16) |
|  | Any Cancer | 1.73 (1.23, 2.44) | 2.01 (0.867, 4.65) | 1.43 (0.869, 2.37) | 1.41 (0.639, 3.1) |
|  | Type 2 Diabetes | 1.91 (1.34, 2.72) | 1.18 (0.488, 2.84) | 2.02 (1.18, 3.45) | 2.79 (1.18, 6.6) |
|  | Kidney | 3.49 (2.39, 5.11) | 2.97 (1.15, 7.66) | 3.68 (2.09, 6.48) | 3 (1.24, 7.27) |
|  | Liver | 1.43 (0.91, 2.26) | 0.595 (0.167, 2.11) | 1.45 (0.735, 2.88) | 2.74 (1.06, 7.06) |
|  | Autoimmune | 1.50 (1.05, 2.13) | 2.07 (0.905, 4.73) | 1.47 (0.856, 2.54) | 1.38 (0.59, 3.22) |
| **ICU (1) vs Not (0)** | | **Full Cohort**  (n0=1136, n1=112) | **Quarter 1**  (n0=203, n1=38) | **Quarter 2**  (n0=312, n1=56) | **Quarter 3**  (n0=347, n1=18) |
| **Age (unit: 10-year)** | | 1.26 (1.11, 1.44) | 1.73 (1.28, 2.34) | 1.04 (0.872, 1.23) | 1.28 (0.959, 1.72) |
| **Age Range**  REF: [18,35) | [0,18) | 2.85 (0.98, 8.34) | NA (NA, NA) | 3.15 (0.828, 12) | 0.662 (0.0335, 13.1) |
|  | [35,50) | 0.75 (0.33, 1.72) | 8.76 (0.524, 147) | 0.216 (0.0597, 0.782) | 1.66 (0.321, 8.56) |
|  | [50,65) | 1.36 (0.67, 2.76) | 18.7 (1.18, 294) | 0.51 (0.195, 1.33) | 0.963 (0.174, 5.33) |
|  | [65,80) | 3.01 (1.46, 6.24) | 32.7 (1.87, 569) | 1.09 (0.421, 2.82) | 5.03 (1.06, 24) |
|  | [80,100) | 3.94 (1.55, 10) | 35.7 (1.79, 714) | 1.45 (0.386, 5.43) | 2.5 (0.184, 33.9) |
| **Male Sex** | | 2.55 (1.69, 3.85) | 2.84 (1.27, 6.36) | 1.98 (1.1, 3.56) | 3.16 (1.14, 8.77) |
| **BMI** | | 1.01 (0.98, 1.04) | 1.02 (0.961, 1.08) | 0.997 (0.957, 1.04) | 1.03 (0.962, 1.11) |
| **BMI Range**  REF: [18.5,25) | <18.5 | 4.18 (0.59, 29.9) | NA (NA, NA) | NA (NA, NA) | 9.55 (0.896, 102) |
|  | [25,30) | 0.96 (0.504, 1.82) | 1.17 (0.336, 4.06) | 1.1 (0.449, 2.68) | 0.798 (0.168, 3.8) |
|  | >=30 | 0.844 (0.452, 1.57) | 0.992 (0.278, 3.54) | 0.667 (0.285, 1.56) | 1.13 (0.256, 5.01) |
| **Ever-Smoker** | | 1.23 (0.803, 1.89) | 2.02 (0.893, 4.58) | 0.76 (0.398, 1.45) | 1.67 (0.61, 4.55) |
| **Smoking Status**  REF: Never-Smoker | Past-Smoker | 1.3 (0.834, 2.02) | 2.07 (0.895, 4.79) | 0.832 (0.424, 1.63) | 1.74 (0.61, 4.95) |
|  | Current-Smoker | 0.899 (0.314, 2.58) | 2.12 (0.279, 16.2) | 0.514 (0.119, 2.22) | 1.76 (0.241, 12.9) |
| **Alcohol Consumption** | | 0.77 (0.49, 1.21) | 1.15 (0.472, 2.81) | 0.692 (0.357, 1.34) | 0.708 (0.244, 2.06) |
| **Race/Ethnicity**  REF: White | Black | 1.11 (0.66, 1.86) | 0.924 (0.353, 2.41) | 0.842 (0.401, 1.77) | 0.321 (0.0489, 2.1) |
|  | Other / Known Ethnicity | 1.09 (0.54, 2.2) | 1.53 (0.43, 5.46) | 0.859 (0.29, 2.55) | 0.969 (0.204, 4.61) |
|  | Other / Unknown Ethnicity | 0.13 (0.01, 2.22) | 0.73 (0.03, 17.6) | 0.248 (0.0126, 4.91) | 0.44 (0.0205, 9.44) |
| **SES** | Population density (1000-people/mi^2^) | 1.11 (1, 1.22) | 1.1 (0.907, 1.33) | 1.1 (0.967, 1.26) | 0.989 (0.78, 1.25) |
|  | NDI | 4.02 (0.334, 48.4) | 34.8 (0.256, 4730) | 0.27 (0.00638, 11.4) | 263 (0.933, 74100) |
| **Comorbidity Score** | | 1.49 (1.28, 1.72) | 1.27 (0.954, 1.68) | 1.49 (1.2, 1.85) | 1.61 (1.14, 2.28) |
| **Comorbidities** | Respiratory | 1.34 (0.741, 2.43) | 1.17 (0.357, 3.82) | 1.32 (0.6, 2.92) | 1.8 (0.442, 7.36) |
|  | Circulatory | 1.76 (0.909, 3.41) | 1.41 (0.455, 4.4) | 3.31 (1.19, 9.23) | 0.747 (0.2, 2.79) |
|  | Any Cancer | 1.83 (1.18, 2.82) | 1.17 (0.479, 2.86) | 2.08 (1.13, 3.84) | 1.24 (0.413, 3.75) |
|  | Type 2 Diabetes | 2.13 (1.37, 3.33) | 1.09 (0.42, 2.83) | 1.83 (0.953, 3.52) | 5.81 (1.98, 17) |
|  | Kidney | 3.82 (2.4, 6.07) | 2.88 (1.12, 7.35) | 4 (2.06, 7.76) | 2.12 (0.686, 6.58) |
|  | Liver | 1.30 (0.73, 2.31) | 0.65 (0.17, 2.51) | 0.797 (0.331, 1.92) | 6.35 (2.07, 19.5) |
|  | Autoimmune | 2.03 (1.3, 3.15) | 3.07 (1.26, 7.52) | 1.88 (0.984, 3.6) | 2.11 (0.699, 6.36) |

Abbreviations: OR, odds ratio; ICU, intensive care unit; BMI, body mass index; NA, not applicable; REF, reference group; SES, social economics status; NDI, 2010 Neighborhood Socioeconomic Disadvantage Index.

Note: green represents significant OR<1 (protective effect), orange represents significant OR>=1 (risk effect) under P<0.05.
